# Longitudinal clinical proteomics reveals pneumonia type-specific protein biomarkers and autoantibodies

**DOI:** 10.64898/2026.01.12.26343938

**Authors:** Anna Semenova, Taylor A. Poor, Johannes B. Müller-Reif, Sai Rama Sridatta Prakki, Phillip Geyer, Martin Mück-Häusl, Lesca M. Holdt, Daniel Teupser, Matthias Mann, Ali Ö. Yildirim, Richard G. Wunderink, Alexander V. Misharin, Ben D. Singer, G.R. Scott Budinger, Theodore S. Kapellos, Herbert B. Schiller

## Abstract

Community-acquired pneumonia is a major cause of morbidity and mortality globally. Specific molecular endotypes are currently not well defined and different viral or bacterial pathogens may trigger specific host responses and pathogenic mechanisms. We performed longitudinal proteomic profiling of bronchoalveolar lavage fluid and plasma from bacterial, influenza and SARS-COV-2 driven pneumonia. Our analysis revealed highly pneumonia type specific proteomic signatures, including COVID-19 specific antibodies locally produced in the lung. These antibodies showed biased immunoglobulin V-domain usage, linked to a *CD69*/*CD83* plasma cell state associated with disease severity and degree of autoimmunity. Using mass spectrometry driven autoantibody profiling in two independent COVID-19 cohorts, we identified 177 putative autoantibodies targeting extracellular matrix, nuclear, and immune-related proteins. Of note, temporal changes in autoantibody profiles correlated with clinical markers of inflammation, organ dysfunction, and duration of hospitalization. These findings highlight the autoimmune aspects of COVID-19 and provide potential biomarkers and therapeutic targets to help improve patient outcomes.

## Introduction

Respiratory infections represent a major global health burden and the leading cause of death from infectious disease worldwide ^1^. Bacterial, viral or fungal infections of the distal airways and alveoli directly damage the lung and induce local and systemic immune responses that present as the clinical syndrome of pneumonia. In severe pneumonia, damage to the alveolar-capillary barrier results in alveolar flooding and the acute respiratory distress syndrome, sometimes complicated by respiratory failure and death ^2^. Severe pneumonia and its complications are more common in the very young, the elderly and immunocompromised ^3^.

The COVID-19 pandemic illustrated the devastating impact of infectious respiratory diseases on healthcare systems and national economies ^4,5^. The large number of patients requiring hospitalization for severe COVID-19 provided an opportunity to understand pneumonia pathobiology and sequelae from the perspective of an individual pathogen and to compare it to pneumonia caused by other pathogens.

SARS-CoV-2 enters the lung through droplet aspiration, where it infects alveolar type 2 cells and alveolar macrophages, and is sustained by inflammatory signaling loops between activated T cells and alveolar macrophages that persists until the virus is cleared ^6,7^. Antigen-specific T cell responses targeting key viral proteins are critical for viral clearance, but ineffective T cell responses drive immunopathology ^8^.

B cell-mediated responses also play an important role in COVID-19 pathobiology. Although antibodies against SARS-CoV-2 play a protective role by blocking viral entry via ACE2 receptor interaction ^9^, dysregulated humoral responses, characterized by extrafollicular B cell activation, low somatic hypermutation ^10^, Fc-mediated proinflammatory cytokine production, and auto-reactive antibodies, exacerbate immunopathology ^11,12^.

Immune dysregulation and autoimmunity may contribute to acute COVID-19 but also play a role in the prolonged sequelae of COVID-19 reported in a substantial number of patients. For example, investigators have reported autoantibodies targeting type I interferons ^12^, ACE2, G protein-coupled receptors, and components of the vascular and coagulation systems ^13–16^ in patients with COVID-19. Autoantibodies have also been reported against nuclear, phospholipid, cytoplasmic, immune signaling molecules, cardiac antigens, and extracellular matrix proteins, however, their clinical relevance remains incompletely understood. Transfer of immunoglobulins leads to post-acute sequelae of COVID-19 (PASC)-like symptoms in mice ^17,18^, while others have been linked to thrombotic complications in acute disease ^15,19^. Intriguingly, autoantibodies against chemokines correlate with protection from PASC ^11^. Many of them also occur in healthy individuals ^20–23^, highlighting a critical gap in our understanding of their pathogenic relevance.

We hereby employed an integrated multi-omics approach to dissect local and systemic immune responses in bronchoalveolar lavage fluid (BALF) and blood samples from patients with COVID-19 compared to patients with pneumonia secondary to other viruses or bacteria. Mass spectrometry detected pneumonia type-specific protein signatures in two independent patient cohorts, several also detectable in plasma. Single-cell RNA sequencing (scRNA-seq) further identified COVID-19-specific immunoglobulin V-segment usage in peripheral blood linked to a distinct plasma cell state, enriched for autoimmune-related genes. Using a differential antigen capture (DAC) assay, we profiled circulating putative autoantibodies, several of which correlated with disease severity and hospitalization length. Collectively, our findings provide a comprehensive framework of pneumonia-associated autoantibodies and protein biomarkers that highlight their potential roles in COVID-19 pathophysiology.

## Results

### Mass spectrometry reveals pneumonia type-specific protein signatures in bronchoalveolar fluid

To identify pneumonia type-specific biomarkers, we sampled bronchoalveolar lavage fluid (BALF) and plasma from individuals enrolled in the Successful Clinical Response in Pneumonia Therapy (SCRIPT) study at Northwestern Memorial Hospital in Chicago (hereafter referred to as the “Chicago cohort”). The cohort encompassed control donors (n=7) and patients requiring mechanical ventilation in the intensive care unit (ICU) who were diagnosed with pneumonia of bacterial (n=6), influenza (n=7) or COVID-19 (n=13) origin (Table S1). Sample collection occurred at the earliest within 48 hours of intubation and collection continued over the entire length of ICU stay, reaching a maximum of 5 time points per patient over up to 60 days after intubation (Fig. 1a-b). Pneumonias of different origins were substantially different in their clinical presentation and host immune responses, as exemplified by the significantly higher (p=0.00011) end-expiratory pressure (PEEP) in COVID-19 and neutrophil relative frequencies (p=0.02) in bacterial pneumonia. In contrast, the sequential organ failure assessment (SOFA) score was similar across groups, suggesting a comparable degree of tissue damage across pneumonia types (Fig. 1c).

**Figure 1.**
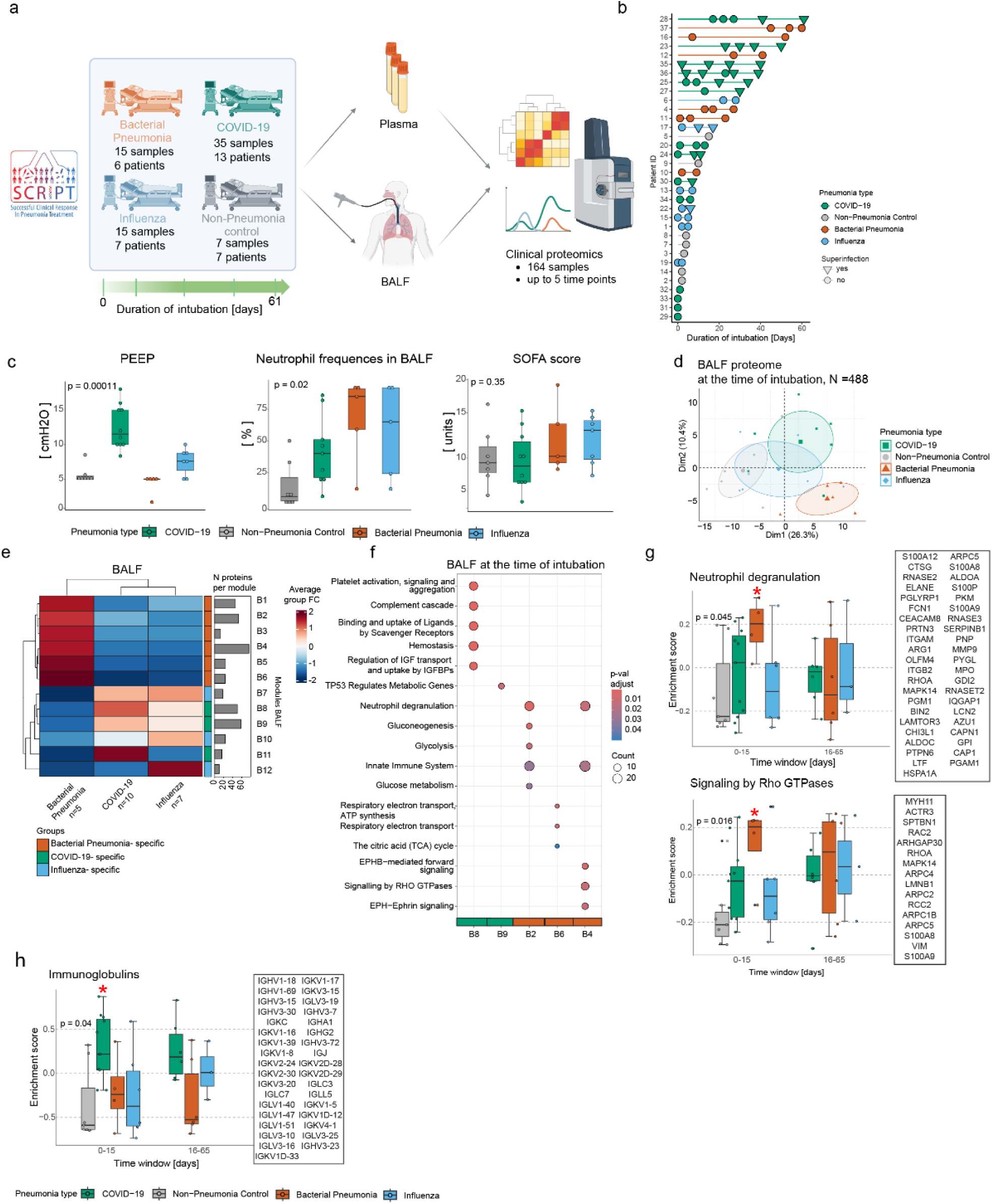
Pneumonia-specific signatures can be detected in BALF upon intubation in the intensive care unit. (a) Overview of study design. (b) Lollipop chart displaying patient numbers and the longitudinal resolution of sample collection per pneumonia type. (c) Box plots that compare clinical parameters and phenotypical observations across pneumonia types. Data are represented as mean ± SD, and variance was statistically assessed with the non-parametric Kruskal-Wallis test. (d) Principal component analysis of BALF proteomes at the time of intubation based on the 488 most variable proteins. (e) Hierarchical clustering of 12 BALF pneumonia-specific protein modules at day 0 of intubation as derived from co-expression network analysis. The color scale denotes the mean fold change of the proteins in each module. (f) Pathway analysis of selected BALF module proteins at intubation induction. The top 5 enriched Reactome terms for each module are displayed. Color code depicts the adjusted p-value, and point size refers to the number of proteins detected per term. (g-h) Box plots displaying the longitudinal enrichment of (g) neutrophil degranulation and (h) immunoglobulin Reactome terms in BALF specimens across pneumonia types. Each dot represents the enrichment score for an individual patient. Data are represented as mean ± SD, and the variability between groups was statistically assessed with the non-parametric Kruskal-Wallis test. Proteins involved in the term are displayed on the right side of the plot.

We next sought to understand how the clinical groups are different on the molecular level. We applied mass spectrometry-based proteomics to BALF specimens from 32 patients, which quantified 985 ± 275 proteins. Principal component analysis (PCA) of the first time-point measurements showed that the proteome profiles of different pneumonia types are well separated already at baseline (Fig. 1d); bacterial pneumonia specimens were distinguished by PC1, whereas COVID-19 pneumonias were separated by PC2. The 411 most variable proteins were then used to construct a co-expression network analysis of the BALF proteomes to identify features that drive the heterogeneity in pneumonia types.

Proteins with similar expression patterns were grouped into modules and were visualized in a heatmap (Fig. 1e). Hierarchical clustering identified 12 protein modules (6 bacterial pneumonia-specific, 3 COVID-19-specific, 3 influenza-specific), which we further interrogated using pathway analysis. Proteins belonging to the bacterial pneumonia-specific modules were enriched, among other biological processes, in neutrophil degranulation (module B2), Signalling by Rho GTPases (module B4), and respiratory electron transport (module B3) (Fig. 1f; Fig. S1a; Table S2). Consistent with these findings, the abundance of neutrophil degranulation proteins correlated positively (R=0.74; p=0.001) with BALF neutrophil frequencies in bacterial pneumonia (Fig. S1b). COVID-19-specific modules were enriched in the regulation of the complement cascade (module B8), platelet degranulation (module B8), p53 metabolic regulation (module B9), and immunoglobulins (modules B8, B9, B11) (Fig. 1f; Fig. S1c; Table S3).

To determine how these proteomic signatures were regulated longitudinally, we also analyzed their enrichment also at later timepoints (Fig. 1g-h; Fig. S1d-e). During the early interval (days 1–15), bacterial pneumonia was uniquely characterized by significant enrichment of neutrophil degranulation and Rho GTPase effector pathways, which lost statistical significance at later time points (Fig. 1g). In contrast, the COVID-19 pneumonia cases showed a pronounced increase in immunoglobulin-related, complement, and platelet activation signatures that remained persistently elevated also at later time points compared to the other groups (Fig. 1h; Fig. S1d-e).

Taken together, we identified BALF protein signatures that distinguish between bacterial, viral and COVID-19 pneumonia types. COVID-19 patients exhibited persistently increased levels of complement cascade proteins, platelet aggregation markers, and a specific set of immunoglobulins.

### A COVID-19-specific set of antibodies locally enriched in the lung versus blood

The BALF proteome can be derived either by local production in the lung or by leakage of plasma factors into the lung due to acute injury and barrier defects. We made use of the fact that we collected matched BALF and plasma samples to analyze the relative enrichment of proteins in the lung versus the blood.

Using mass spectrometry, we quantified 374 ± 33 proteins in the plasma samples. The PCA of the baseline timepoint measurements revealed a clear separation of the COVID-19 proteomes by PC1 and PC2 (Fig. 2a). Similarly to BALF, co-expression network analysis of the plasma proteomes grouped the proteins in 7 modules (2 bacterial pneumonia-specific, 2 COVID-19-specific, 3 influenza-specific) (Fig. 2b; Table S4). Proteins belonging to the bacterial pneumonia-specific modules were enriched in the regulation of insulin-like growth factor transport and uptake (module S3) (Fig. 2c). COVID-19-specific modules included MAP/K family signaling cascades (module S1) and immunoglobulins (module S2). Lastly, influenza-specific modules displayed strong immune responses related to complement cascades (module S5), platelet activation (module S6), and mRNA splicing (module S7) (Fig. 2c; Fig. S2a; Table S5).

**Figure 2.**
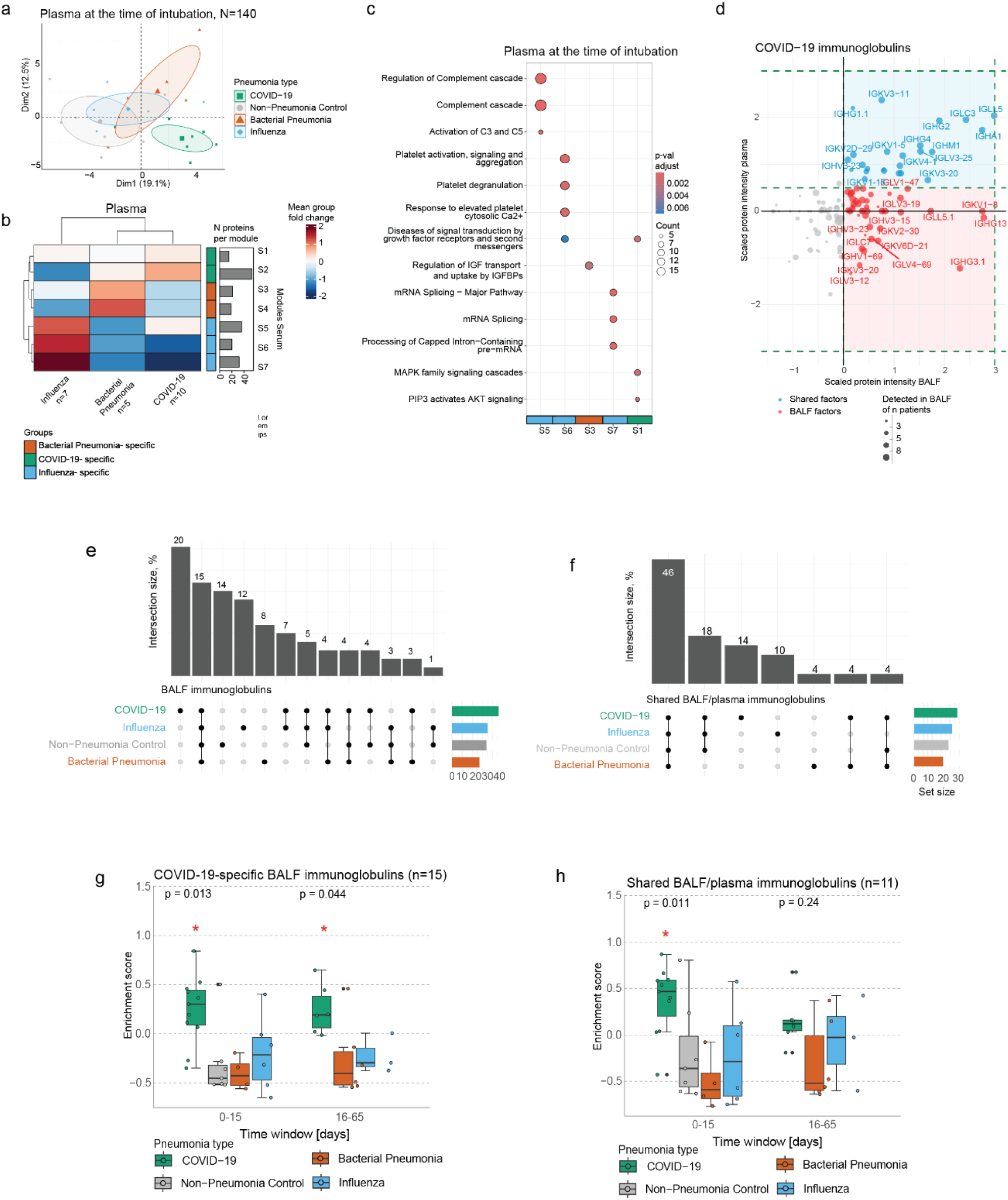
Integrative analysis of body fluid proteomes reveals lung-resident biomarkers. (a) Principal component analysis of plasma proteomes at the time of intubation induction based on the 140 most variable proteins. (b) Hierarchical clustering of 7 plasma pneumonia-specific protein modules at day 0 of intubation as derived from co-expression network analysis. The color scale denotes the mean fold change of the proteins in each module. (c) Pathway analysis of selected plasma module proteins at intubation induction. The top 5 enriched Reactome terms for each module are displayed. Color code depicts the adjusted p-value, and point size refers to the number of proteins detected per term. (d) Scatter plot displaying immunoglobulin segment abundance in the BALF (x-axis) and plasma (y-axis) of COVID-19 patients at the time point upon intubation. Dot size corresponds to the number of patients that express a detected protein. (e-f) Upset plot displaying the overlap of (e) BALF and (f) shared BALF/plasma immunoglobulin segments across pneumonia types. (g-h) Box plots displaying the longitudinal enrichment of COVID-19-specific (g) BALF and (f) shared BALF/plasma immunoglobulin signatures across pneumonia types. Each dot represents the enrichment score for an individual patient. Data are represented as mean ± SD and were statistically assessed with the non-parametric Kruskal-Wallis test.

In both the BALF and plasma, we found COVID-19-specific enrichment of immunoglobulins. Since we measured both biofluids from matched donors, we were able to quantitatively compare the enrichment of proteins that were detected in both the BALF and the plasma (Fig. 2d). Indeed, in the quadrant that represents proteins with high abundance in both compartments, we found a set of antibodies (V-segments) that were highly abundant in both BALF and plasma (highlighted in blue) (Fig. 2d). In addition, several antibodies were quantitatively enriched in BALF and hence likely produced locally (highlighted in red). Interestingly, many locally produced V-segments were highly specific to COVID-19 (20%), influenza (12%) or bacterial pneumonia (8%) (Fig. 2e), in contrast to the systemic V-segments, which were less specific to individual pneumonia (COVID-19 (14%), influenza (10%), bacterial pneumonia (4%)) types (Fig. 2f). Longitudinal assessment of immunoglobulin profiles across pneumonia types confirmed the higher enrichment of COVID-19-specific BALF and shared BALF/plasma segments, evident at both time intervals and significantly pronounced at days 0-15 (Fig. 2g-h).

In summary, our plasma proteomics analysis identified distinct molecular signatures across pneumonia types. In COVID-19, immunoglobulin segments were enriched in both BALF and circulation, exhibiting specificity and longitudinal persistence.

### Clonal expansion of a specific set of plasma cells in severe COVID-19

We next verified the observed COVID-19-specific antibody V-segment bias in an independent multimodal scRNA-seq dataset of peripheral blood samples ^24^, encompassing healthy controls (n=23), as well as patients representing asymptomatic (n=9), mild (n=23), moderate (n=30), severe (n=13), and critical (n=15) stages of COVID-19 (Fig. 3a) ^24^. We re-clustered a total of 38,063 B cells and plasma cells (Fig. S3a), with B cells forming six distinct clusters upon consideration of both their transcriptomes (Fig. S3b) and surface proteomes (Fig. S3c-d). Annotation of the resulting clusters using canonical marker genes ^25^ identified memory, immature, naive and natural B1 cells, as well as dividing plasmablasts and IgA-high plasma cells (Fig. 3b; Table S6). Notably, we observed an increase in the IgG and IgA relative proportion in B/plasma cells from severe disease (Fig. S3e) which was reflected in the significant (p=0.01) increase in the relative frequencies of IgA-high plasma cells (p=6.6 x 10e-6) and dividing plasmablasts across COVID-19 stages. In contrast, naive (p=0.00011) and memory B cells (p=6.2 x 10e-5) displayed the opposite pattern (Fig. S3f).

**Figure 3.**
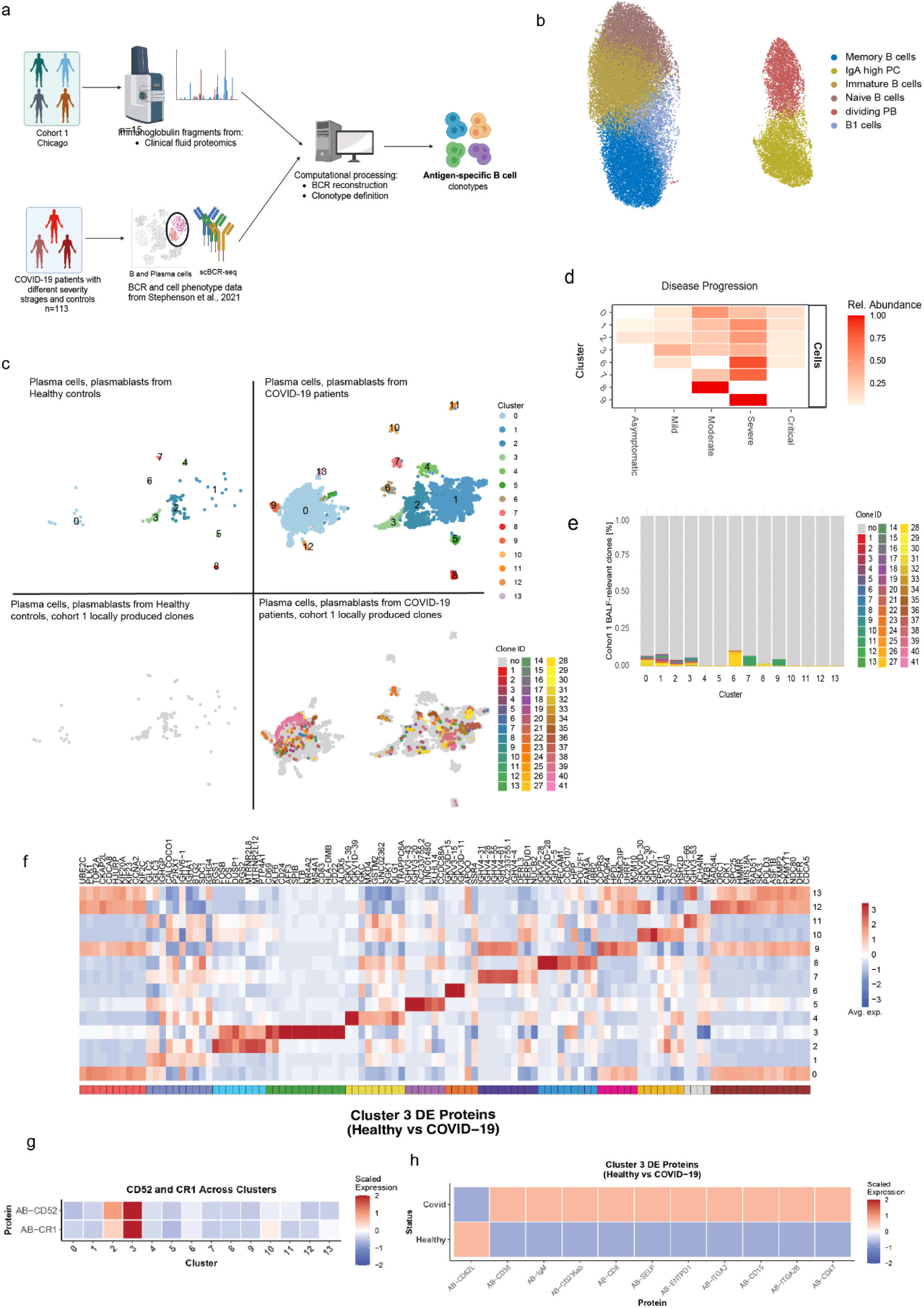
Peripheral B cell clonal dynamics and transcriptional signatures are associated with COVID-19 severity. (a) Overview of bioinformatics workflow. (b) UMAP visualization of 38,063 peripheral blood B/plasma cells from 90 COVID-19 patients and 23 healthy controls, colored by cluster identity. (c) UMAP projection of 8,599 plasma cells from 90 COVID-19 patients and 23 healthy controls. Lower panels show clonal expansions of BALF-derived V-fragment lineages mapped to COVID-19 patients vs controls. (d) Heatmap visualizing the relative abundance of B/plasma cell molecular states across stages of COVID-19 disease progression. (e) Bar plot showing clonal percentage of proteomics-identified V-segments per plasma cell state. (f) Heatmap of top differentially expressed genes for each plasma cell state. Color code denotes the scaled average expression across clusters. (g) Heatmap of top differentially expressed proteins for plasma cell state 3. Color code denotes the scaled average expression across clusters. (h) Heatmap of differentially expressed surface protein markers in plasma cell state 3 between healthy controls and COVID-19 donors.

BCR-seq analysis revealed that the COVID-19-specific V-segment clonotypes identified in our proteomics dataset were predominantly expressed by plasma cells (Fig. S3g). To explore this further, we re-clustered these cells into 14 transcriptomically distinct molecular states (Fig. 3c; Table S7). These plasma cell states were the main source of BALF-derived V-fragments, highlighting their potential role in shaping COVID-19-specific humoral responses. The abundance of plasma states 0-3 progressively increased with severity with advancing disease severity, while others (cluster 6-9) were enriched in moderate and severe cases (Fig. 3d). Notably, these plasma cell states were also the main source of the COVID-19-specific BALF immunoglobulins, highlighting their potential role in shaping disease-relevant humoral responses (Fig. 3e).

Our findings prompted us to perform differential gene expression analysis to reveal how these states are different from each other (Fig. 3f; Table S7). As expected, light and heavy chain V-segments distinguished several plasma cell states. Plasma cell states 1 and 13 were characterized by the high expression of proliferation markers, whereas molecular state 3 represented activated plasma cells, defined by high expression of *CD69* and *CD83*. This plasma cell state has previously been marked by the expression of genes implicated in autoimmune diseases ^26,27^. For instance, *NR4A2* and *LTB* drive the formation of ectopic lymphoid structures and pathological B cell responses in rheumatoid arthritis ^27^, while *AFF3* is a susceptibility factor for RA and type I diabetes ^28^. Additionally, the surface proteins such as *CD52* and *CR1*, which were upregulated in cluster 3, are also implicated in autoantibody production in B cell subsets from patients with rheumatoid arthritis and systemic lupus erythematosus (Fig. 3g; Table S8) ^29,30^. Lastly, analysis of the plasma cell state 3 proteome showed an upregulation of markers, including CD38, ITGA2, ITGA2B, and SELP, suggesting enhanced interactions with the extracellular matrix and the vascular endothelium, as well as localized immune responses (Fig. 3h; Table S8).

Conclusively, we identified transcriptomically distinct plasma cell states that expand with COVID-19 severity and serve as the predominant source of the disease-specific antibody V-segments detected by proteomics. Notably, one of these states exhibited an activated, autoimmune-like transcriptional profile, and featured genes implicated in ectopic lymphoid formation and autoantibody production.

### A differential autoantigen capture assay reveals COVID-19-specific autoantibodies

Given the mounting evidence for autoantibodies in acute COVID-19, as well as PASC, and our observation that severe COVID-19 is associated with a specific V-segment bias, which could favour autoimmunity, we were interested in profiling autoreactive antibodies in our cohort. We recently developed a Differential Autoantigen Capture (DAC) assay ^31^, which is based on immunoprecipitation of natively extracted human lung proteins using patient antibodies captured from their plasma or serum (Fig. 4a). Mass spectrometry quantifies the relative antibody-mediated enrichment of proteins in each patient compared to the average in a control population. Thus, we used the DAC assay to analyze antibody-mediated autoreactivities in the Chicago cohort alongside a second longitudinal cohort of COVID-19 patients (hereafter referred to as the “Munich cohort”), which included mild cases (n=7) and severe ICU-admitted COVID-19 patients (n=16) (Fig. 4b; Table S9). The Munich cohort also included two COVID-19 negative control groups, categorized as low inflammatory (n=24) and high inflammatory (n=23) based on the circulating levels of CRP, creatinine, and leukocytes (Fig. S4a). Importantly, BALF and shared BALF/plasma V-segments were significantly enriched over time in severe COVID-19 patients compared to mild cases and the controls, substantiating the presence and persistence of pneumonia-specific immunoglobulins throughout disease progression (Fig. S4b-c).

**Figure 4.**
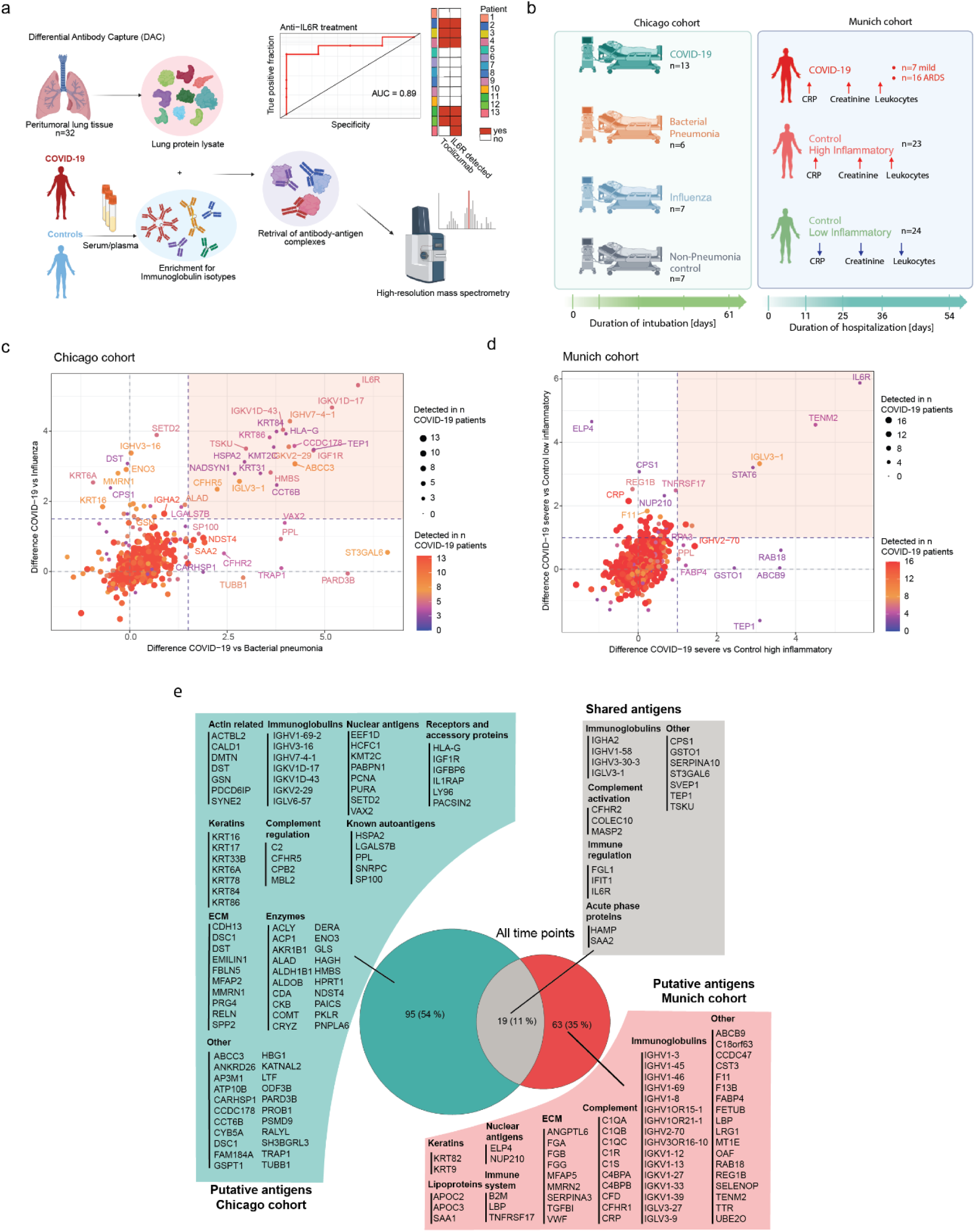
Comparative analysis of the putative auto-antigen repertoire of two independent COVID-19 cohorts. (a) Flowchart of the Differential Antibody Capture assay (DAC) and benchmarking processes with anti-IL-6R antibody. (b) Schema of Chicago and Munich cohort demographics. (c) Scatter plot displaying fold changes of individual antigens in COVID-19 patients (n=13) of the Chicago cohort versus bacterial pneumonia patients (n=6) on the x-axis and influenza patients (n=7) on the y-axis at the first time point upon intubation. Dot size and color represent the number of COVID-19 patients with a particular plasma autoantibody. Enriched putative autoantibodies in COVID-19 over bacterial pneumonia and influenza are depicted in the highlighted rectangle. (d) Scatter plot displaying fold changes of individual antigens in severe COVID-19 patients (n=16) of the Munich cohort versus high-inflammatory control patients (n=23) on the x-axis and low-inflammatory control patients (n=24) on the y-axis at the first point upon in hospitalization. Dot size and color and color represent the number of COVID-19 patients with a particular plasma autoantibody. Enriched putative autoantibodies in COVID-19 over both controls are depicted in the highlighted rectangle. (e) Venn diagram demonstrating putative autoantibody repertoires of the Chicago and Munich cohorts grouped by molecular function. Displayed autoantigens are significantly enriched at least at one time point over at least one control group and in at least 3 COVID-19 patients.

In our previous study, the DAC assay reached almost 100% sensitivity and specificity against a clinical ELISA benchmark ^31^. To test these performance metrics in the present study, we made use of the fact that some patients were treated with the anti-IL6R therapeutic monoclonal antibody Tocilizumab ^32^. We observed that IL6R was significantly enriched over controls only in patients who received the therapeutic antibody (AUC=0.89) (Fig. 4a), demonstrating the high specificity of the immunoprecipitation. To identify putative COVID-19-specific autoantigens in severe disease, patients in the Chicago cohort and severe cases from the Munich cohort were screened, and the enriched antigens in each cohort at the first time point were further analyzed (Fig. 4c-d; Fig. S4d-e). In both cohorts, we found antigens significantly enriched over other pneumonia types (Fig. 4c; Fig. S4d) or controls (Fig. 4d; Fig. S4e) already upon hospitalization, with a prevalence ranging from 3-8 patients across all putative autoantigens.

Overall, we identified 95 putative autoantibodies in the Chicago cohort and 63 putative autoantibodies in the Munich cohort ranging functions from extracellular matrix proteins, keratins, lipoproteins, complement cascade members, nuclear antigens, enzymes, receptors, acute phase proteins and signal transduction proteins (Fig. 4e; Table S10). Several of these antibodies have been associated with autoimmune disease. For example, PPL autoantibodies were described in pulmonary fibrosis donors ^33^, anti-U1 snRNP (SNRPC) have been linked to mixed connective tissue disease and SLE ^34^, whereas anti-SP100 are associated with primary biliary cholangitis ^35^. Despite the substantial inter-individual variance and significant differences between the Chicago and Munich cohorts, we also identified 19 hits (15 putative autoantigens, 4 immunoglobulins) that were shared between cohorts and significantly enriched in at least at one time point over at least one control group and in at least 3 COVID-19 patients with severe disease (Fig. 4e). Among the shared hits, autoantibodies against acute phase response proteins, complement and anti-glutathione S-transferase omega-1 (GSTO1) have been described in autoimmune diseases, SLE, RA and COVID-19, leading to excessive organ damage ^36,37,38^. Notably, the discovery of IFIT1 autoantibodies in our data supports previous work that has described autoantibodies interfering with IFN signaling in severe COVID-19 patients ^12,39^.

In conclusion, we detected antibodies with an affinity for native lung proteins in the plasma of COVID-19 patients from two independent cohorts, suggesting the presence of multiple putative autoantigens in severe COVID-19. Many of these shared and cohort-specific antibodies encompass extracellular matrix proteins, immunoglobulins, complement cascade members, acute phase proteins, and others, highlighting both the diversity and clinical relevance of COVID-19-associated autoimmunity and underscoring the value of unbiased proteomics in revealing novel biomarkers for predicting disease severity.

### Temporal and severity-associated dynamics in autoantibody responses

Since the Chicago cohort encompassed severe COVID-19 samples, we set out to compare the longitudinal profiles in both the severe and mild cases of the Munich cohort. We first performed patient clustering based on all available clinical parameters. This revealed two patient groups; cluster 1 comprised individuals with severe disease manifestations, including acute respiratory distress syndrome (ARDS), immunosuppression, a higher incidence of kidney failure, and increased mortality, whereas cluster 2 included patients with milder symptoms (Fig. 5a). These clinical differences were mirrored in the plasma levels of IL-6 and CRP, biomarkers of systemic inflammation (Fig. S5a). To characterize temporal variations in autoantibody abundance, we subsequently grouped the observations into 3 time windows (days 0-11, 12-36 and 37-56) (Fig. S5b) and applied a mixed-effects linear model, with time and severity as fixed effects and patient ID as a random effect. The generated model identified 52 autoantigens with significant associations with one or both fixed effects (Fig. 5b).

**Figure 5.**
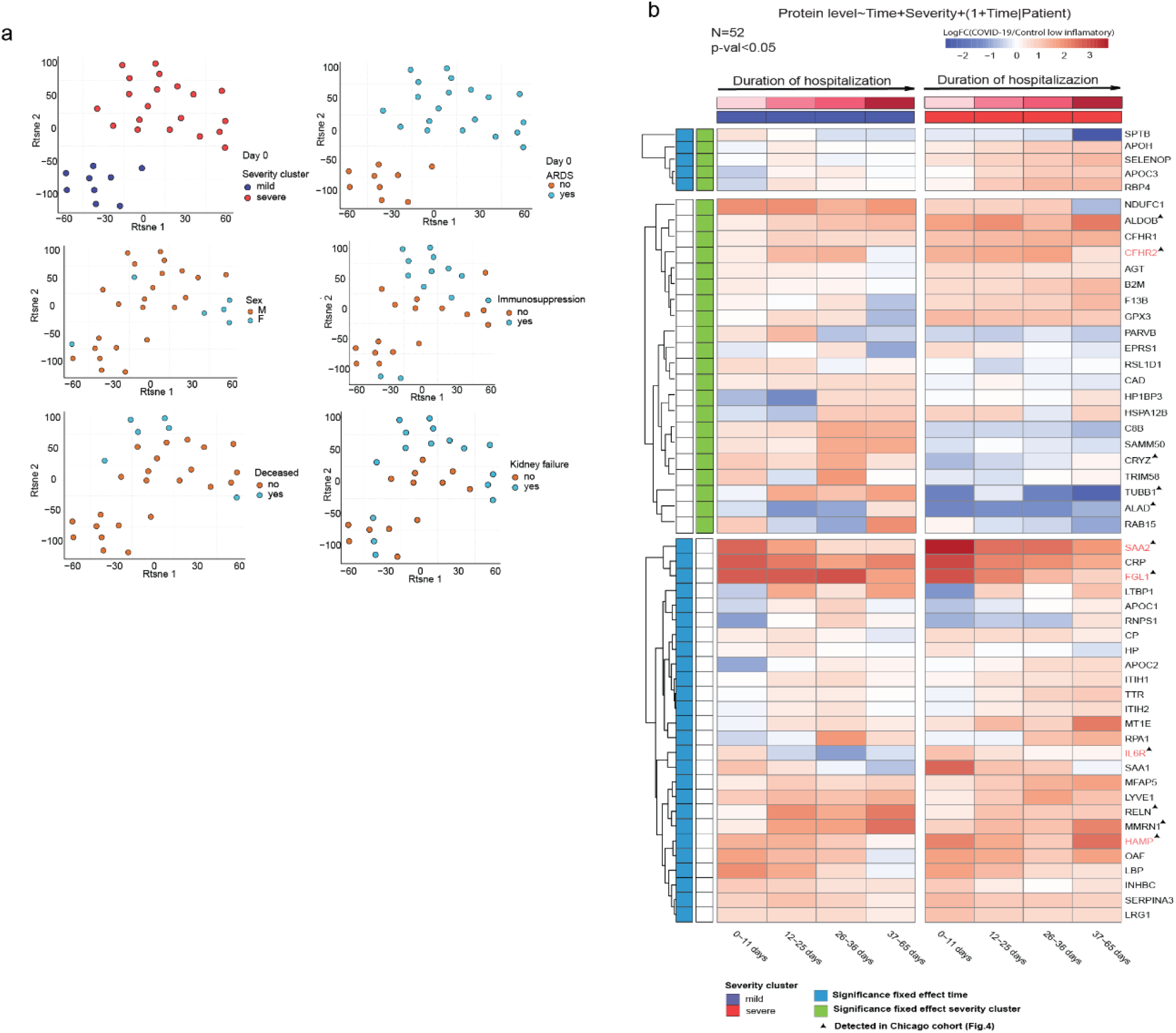
Longitudinal profiling of autoreactivities in mild and severe COVID-19 patients. (a) T-distributed stochastic neighbor embedding (t-SNE) of 27 COVID-19 patients enrolled in the Ludwig-Maximilians-University Hospital (Munich cohort) based on available clinical parameters at the beginning of intubation. Two clusters (mild and severe) are shown. (b) Heatmap of mixed-effects model for time and COVID-19 severity. Only significant proteins (p<0.05) are displayed, and shared proteins with the Chicago cohort are highlighted. Color code represents average expression in time intervals after hospitalization.

Among the identified autoantigens, 5 were significantly associated with both time and severity. In particular, antibodies against APOH, SELENOP, APOC3, and RBP4 were prevalent in severe COVID-19 patients, and their levels increased during hospitalization. Within the putative autoantibodies that were associated with disease severity, severe patients had elevated levels of anti-F13b, anti-B2M, anti-AGT, and anti-GPX3 antibodies compared to mild cases, whereas mild patients had higher levels of anti-TUBB1, anti-ALAD, anti-RAB15, anti-TRIM58, anti-CRYZ, anti-CD8B, anti-SAMM50, anti-PARVB, anti-CAD, ant-RSL1D1, and anti-EPRS1 antibodies (Fig. 5b). Lastly, we detected putative autoantigens that were associated with hospitalization duration in both mild and severe patients. Notably, antigens that were correlated in both severity groups (SAA1, SAA2, LBP, FGL1) were related to acute inflammation and decreased over time. In contrast, those correlating in severe cases (IL6R, CRP) showed both upward and downward trends over time (Fig. 5b).

Taken together, we followed the dynamics of putative autoantibodies in the circulation of a longitudinal cohort of COVID-19 patients and we described immunoglobulins that display temporal or severity-related abundance patterns.

### Autoreactivities correlate with clinical parameters and severity in COVID-19 patients

We subsequently assessed the abundance of the identified autoreactivities related to key clinical features that included measures of lung function (P/F ratio, plateau pressure, PEEP, SOFA score), inflammation and anti-viral immunity (CRP, ferritin, procalcitonin, IL-6, anti-N antibodies, anti-S antibodies), coagulation and thrombosis (D-dimer, fibrinogen, platelet counts), multi-organ injury (ALT, AST, LDH, troponin I, troponin T, creatinine, creatine kinase), disease severity and prognosis (maximum COVID stage, cumulative ICU days, intubation days, BMI, age) and leukocyte composition (neutrophil, macrophage, lymphocyte counts). In both cohorts, we identified a broad spectrum of associations between the putative autoantibodies and clinical parameters upon admission to the hospital. The assessment of associations between the shared autoantibodies in the Chicago and Munich cohorts and clinical parameters revealed a few putative associations with the number of days in intubation, coagulation, liver enzymes, anti-viral inflammation, as well as blood leukocyte and platelet abundance (Fig. 6a-b).

**Figure 6.**
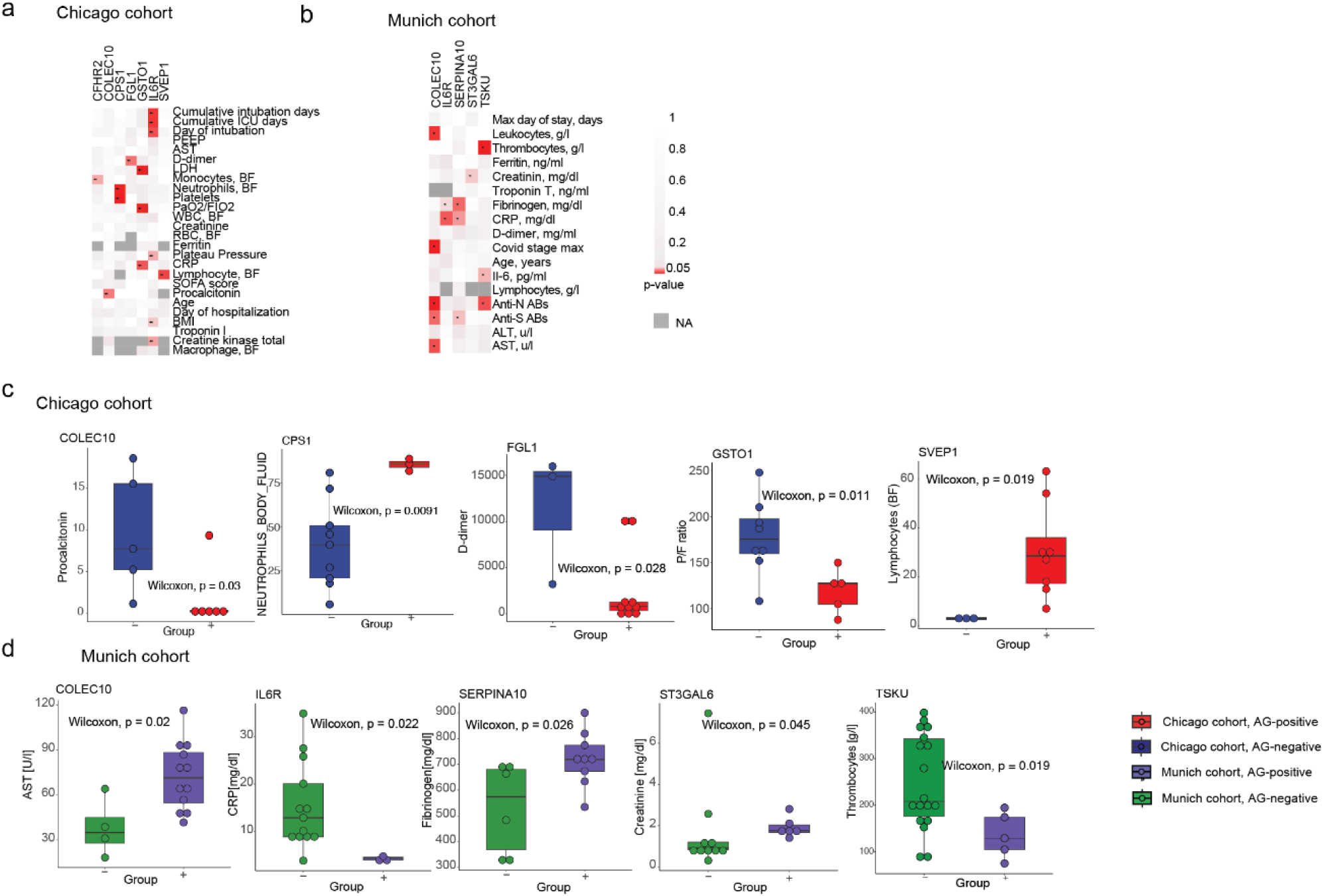
Blood putative autoantibodies are associated with clinical parameters in two independent severe COVID-19 cohorts. (a-b) Heatmaps presenting associations between the detection antibodies against shared putative antigens (n=15) in the peripheral blood of (a) n=13 COVID-19 patients of the Chicago and (b) n=16 severe COVID-19 patients of the Munich cohort upon intubation on the x-axis and selected clinical parameters on the y-axis in the Chicago and Munich cohorts. The color denotes the output of the Wilcoxon test (p-value). The dark grey color corresponds to the absence of the test results due to missing values. (c-d) Box plots showing the significant associations between 19 putative shared autoantigens and clinical parameters in two cohorts: Chicago (c) and Munich (d). The x-axis represents COVID-19 patients categorized based on the presence (+) or absence (-) of detected autoantigens at the time of intubation. Statistical significance was assessed using the Wilcoxon test.

In both cohorts, we observed a broad range of associations between the putative autoantigens and clinical parameters at the time of hospital admission (Fig. 6a-b). Some of the most prominent shared patterns linked autoreactivities to prolonged intubation, coagulation abnormalities, elevated liver enzymes, anti-viral responses and altered blood cell counts. Within the shared autoantigens, several targets (IL6R, TSKU, CPS1, FGL1, SVEP1, CFHR2, ST3GAL6, GSTO1, SERPINA10, COLEC10) demonstrated correlations, spanning respiratory compromise (PaO_2_/FiO_2_ ratio, plateau pressure, D-dimers), systemic inflammation (IL-6, CRP, procalcitonin), coagulation activation (fibrinogen, platelet counts), tissue injury (LDH, AST, creatinine, creatine kinase), clinical severity (maximum COVID-19 stage, BMI, intubation duration), antibody responses (anti-N, anti-S) and immune cell counts (leukocytes, monocytes, neutrophils, lymphocytes).

In addition, we detected cohort-specific patterns. In the Chicago cohort, autoreactivities were directed, among others, at lung structural proteins (KRT6A, KRT78, KRT84, KRT86), desmosomes (DST, PPL, DSC1), the actin cap (GSN, CALD1, ACTBL2, HCFC1), and meiotic synapses (HSPA2, SYNE2). These autoantibody profiles were associated with patient outcomes, encompassing respiratory function (PaO_2_/FiO_2_ ratio, PEEP, plateau pressure), markers of inflammation and tissue injury (procalcitonin, LDH, creatine kinase, D-dimers), cell frequencies (lymphocyte, platelet and total blood cell counts), and disease burden (intubation, duration of hospitalization, ICU stay, BMI, age) (Fig. S6a). In the Munich cohort, distinct autoreactivities targeting proteins, including ABCB9, RAB18, REG1B, NUP210, and ANGPTL6 were associated with changes in thrombocyte counts, ferritin and creatinine levels, the maximum COVID-19 stage, D-dimer and IL-6 levels, and anti-viral antibody titers (Fig. S6b).

To test whether the presence of shared autoantibodies was associated with differences in clinical status, we compared the values of the aforementioned clinical parameters, including indicators of lung function, systemic inflammation, coagulation activity and multi-organ injury, between patients positive and negative for the shared autoreactivities across both cohorts, and performed the same for the cohort-specific autoreactivities (Fig. 6c-d; Fig. S6c-d). This analysis showed that antibody-positive patients often displayed more severe impairments in, and higher indices of disease severity than antibody-negative patients, reinforcing the link between specific autoantibody profiles and the clinical manifestations of severe COVID-19.

To summarize, our analysis revealed clear associations between the levels of circulating autoantibodies and multiple clinical parameters in COVID-19 patients. Based on the biological functions of their target antigens, the detected autoantibodies may disrupt key molecular pathways, contributing to multi-organ injury and coagulopathy, commonly observed in severe cases. Moreover, their presence may prolong intubation and hospitalization requirements and increase susceptibility to secondary bacterial infections.

## Discussion

The specificity of cellular and molecular processes and the associated biomarkers in COVID-19 compared to the sequelae of other severe respiratory infections is not yet well established. Our study builds a solid framework of COVID-19-specific biomarkers in comparison to influenza and bacterial-mediated pneumonia using longitudinal proteomic profiling of both BALF and blood. In particular, we identified a COVID-19-specific set of plasma cell clonotypes and their corresponding antibodies that may contribute to the observed increase in B cell-mediated autoimmunity in severe COVID-19 and possibly its long-term sequelae (long-COVID). We found that autoantibody abundance correlated with both hospitalization duration and disease severity, and was further associated with critical clinical indicators, including lung function decline, disease progression, leukocyte counts, coagulation abnormalities, systemic inflammation, and multi-organ failure. Whether these autoantibodies are pathogenic or biomarkers of disease severity will require prospective validation.

As expected, we observed distinct differences in the clinical presentation between pneumonia types. These clinical patterns were mirrored in pneumonia type-specific biological pathways ^40,41^. Importantly, these proteomic patterns persisted throughout the patient intubation period, underscoring the distinct pathophysiological mechanisms governing each pneumonia type. Importantly, key biological processes were consistently detected in both BALF and plasma, suggesting that loss of endothelial barrier integrity allows lung-derived proteins to leak into the circulation, thereby promoting inflammation and obstructing recovery ^42–45^. This highlights the potential of these blood proteomic signatures as clinically relevant biomarkers.

Humoral responses, an essential component of the adaptive immune system, are responsible for recognizing and neutralizing external threats. In healthy individuals, both naive and memory B cell compartments may harbor autoreactive clones under tight regulatory control. However, in severe inflammatory conditions, such as in COVID-19, SLE, and other autoimmune disorders, autoreactive, age-associated, and atypical B cell populations can expand and secrete pathogenic autoantibodies ^46–50^. Following SARS-CoV-2 infection, B cells initiate IgM production, followed by IgG and IgA responses that can persist for over a year ^51,52^. However, in severe cases, germinal center formation is impaired, promoting extrafollicular B cell responses that favor rapid differentiation into plasma cells ^10,53^ and the production of low-affinity, broad autoreactive antibodies. These antibodies often target nuclear antigens, phospholipids, and carbamylated proteins ^16,25,54^ and are linked to poor clinical outcomes ^16^. While autoreactive plasma cells typically contract during convalescence, serological autoreactivity can persist in some patients and may contribute to PASC ^55,56^. In particular, antibodies with altered glycosylation or fucosylation patterns can bind to Fcγ receptors on myeloid cells, triggering excessive pro-inflammatory cytokine release ^57^. Lastly, cross-reactive B cell clonotypes that recognize both viral and self-antigens may play a dual role in antiviral defense and autoimmunity ^58^.

We found that the abundance of immunoglobulin fragments in COVID-19 BALF was increased across severity stages, and we identified a transcriptionally distinct population of activated plasma cells that expressed a broad repertoire of these proteomics-matched clonotypes. These cells expressed *CD69* and *CD83* and showed transcriptomic signatures linked to autoimmune diseases such as rheumatoid arthritis, type I diabetes, and SLE. CD69 engagement has been linked to TGF-β production, whereas its deficiency exacerbated inflammation in a murine collagen-induced model of arthritis ^59^, suggesting a dual role in regulating inflammation. This population may therefore constitute a putative target to mitigate autoimmunity progression without compromising antiviral immunity ^47^, while whether it could serve as a predictive biomarker for post-infectious autoimmunity in PASC remains to be determined and should be the focus of future longitudinal studies.

The discovery of plasma cell molecular states associated with autoimmunity prompted us to apply our novel mass spectrometry-based DAC assay, which allows unbiased and multiplexed identification of antibody-mediated autoreactivities ^31^. Using a mixed-effects model to disentangle the effects of disease severity and intubation time, we identified autoantibodies that target components of the type I interferon pathway ^12^, as well as anti-phospholipid antibodies, also implicated in aberrant coagulation in a mouse model ^60^. Additionally, we found autoantibodies against cytokines and other immune signaling molecules ^39^. Extending previous findings, we detected autoantibodies directed against C1q, β2GP1, SERPINA10, and SVEP1, which are linked to thrombosis, hypercoagulability, and microvascular damage, commonly observed in severe COVID-19 ^60–62^. We also measured autoantibodies against intracellular proteins such as CPS1, ST3GAL6, GSTO1, ABCB9, RAB18, and NUP210 or epithelium lining components (KRT6A, KRT78, KRT84, KRT86), suggesting that these auto-reactivities may derive from excessive tissue damage. Of note, we found that the levels of several autoantibodies correlated with clinical markers of organ damage, such as elevated CRP, serum creatinine, liver transaminases, and respiratory dysfunction, pointing to a possible role as either drivers or a consequence of multi-organ dysfunction. Hence, our findings expand our understanding of COVID-19-associated autoreactivities and highlight how a diverse array of emerging cross-reactive antibodies may shape the clinical course, severity, and multi-organ failure, hallmarks of severe disease.

Our study has certain limitations. First, differences in sample procurement between cohorts may have introduced bias. The Chicago cohort was enriched for severe COVID-19 cases as enrollment in the cohort required BAL fluid sampling and included paired plasma samples, whereas the Munich cohort primarily analyzed serum. Additionally, both cohorts were relatively small, which may limit the generalizability of our findings, particularly given the high heterogeneity of autoantibody targets and the low detection frequency of specific autoantibodies in severe COVID-19 patients ^63,39^. Second, the sensitivity of the DAC assay is influenced by several factors, including low antibody titers or affinity in patient plasma, as well as the abundance and accessibility of autoantigens within the lung tissue lysate, potentially leading to missed autoantibody specificities. Moreover, our focus on lung-derived antigens may have overlooked autoantibodies targeting other organs. Finally, we were unable to assess pre-infection autoantibody levels in our cohorts, limiting our ability to distinguish between pre-existing autoantibodies and those induced by SARS-CoV-2 infection.

In conclusion, our data have direct implications for enhancing the clinical management of severe COVID-19. While earlier studies have proposed an autoimmune component, our work strengthens this notion by linking distinct autoantibody signatures to molecularly phenotyped plasma cell populations and defined clinical features. These autoantibodies may not only serve as biomarkers for disease monitoring but also as potential therapeutic targets in patients with PASC. Future studies should explore how modulating the autoreactive immune landscape might mitigate long-term complications and refine personalized treatment strategies in COVID-19 and other post-infectious syndromes.

## Methods

### Chicago cohort

Samples from COVID-19, bacterial pneumonia, influenza, and non-pneumonia control patients were collected from participants enrolled in the Successful Clinical Response in Pneumonia Therapy (SCRIPT) study (STU00204868) ^6^. Participants were adults with clinical suspicion of pneumonia, indicated by fever, radiographic infiltrates, and respiratory secretions. All patients experienced respiratory failure requiring mechanical ventilation in the ICU. Intubation decisions were based on bedside clinicians’ assessments of worsening hypoxemia, hypercapnia, or inadequate response to high-flow oxygen or non-invasive ventilation. Extubation decisions followed a protocol-guided evaluation of spontaneous breathing and cardiorespiratory improvement. Critical care physicians at Northwestern Memorial Hospital retrospectively classified patients into five groups: COVID-19 pneumonia, non-COVID-19 viral pneumonia, pneumonia from other pathogens, non-pneumonia controls, and a control group, according to a standardized adjudication process.

BAL fluid was screened for methicillin-resistant *Staphylococcus aureus* (MRSA) using PCR (MRSA/SA SSTI), the BioFire FilmArray Respiratory 2 (RP2) panel, and Pneumonia panels. Clinical laboratory data were obtained from the Northwestern Medicine Enterprise Data Warehouse. To focus on pneumonia type specific signatures, samples from viral pneumonia patients (COVID-19, influenza) with bacterial superinfections at later time points were excluded from the initial analysis. This filtering resulted in 10 COVID-19 patients, 7 influenza patients, 6 bacterial pneumonia patients, and 7 non-pneumonia control patients (Fig. 1e; Table S1). The clinical subgroups were matched for age, race, and sex, exhibiting diverse clinical outcomes.

### Munich cohort

Serum samples of SARS-CoV-2 positive patients collected longitudinally between March and June 2020 at the Ludwig Maximilian University of Munich Hospital (LMU) ^64^. Sampling extended up to 54 days post-admission and included patients treated in both regular wards and intensive care units (ICU). All serum samples were stored at −80°C in the LMU LabMed Biobank.

### Sample collection

Bronchoscopy, often accompanied by blood draws, was performed as part of routine clinical care to guide antimicrobial therapy. Bronchoalveolar lavage fluid (BALF) and plasma samples from participants in the SCRIPT study were collected between June 15, 2018, and July 6, 2020, in the ICU at Northwestern Memorial Hospital, Chicago. Clinical laboratory testing followed standard ICU protocols and included multiplex PCR (BioFire Film Array Respiratory 2 panel), automated cell counts, and urinary antigen testing for *Streptococcus pneumoniae* and *Legionella pneumophila* serogroup 1, all performed on the day of admission. For intubated ICU patients, BALF was collected using single-use aScope devices (Ambu, USA) under sedation and topical anesthesia. The bronchoscope was wedged into the lung area of interest based on chest imaging or procedural observations. Sequential aliquots of 30 ml normal saline were instilled and aspirated, totaling 90 to 120 ml. The fluid recovered from the first aliquot was discarded. BAL samples were split between clinical diagnostics and research. Non-bronchoscopic BALF (NBBALF) followed a similar procedure but was performed by a respiratory therapist using directional guidance rather than by a pulmonologist.

For COVID-19 patients, sampling targeted the lung region with the greatest radiographic abnormality and was performed by a critical care physician using a disposable device. Sedation and neuromuscular blockade were administered to prevent coughing during bronchoscopy. The earliest BALF procedures were conducted immediately post-intubation to utilize the existing neuromuscular blockade.

Whole blood was collected from SCRIPT cohort patients in lithium heparin tubes on the same day as BALF or NBBALF. Plasma was separated by centrifugation at 1,690 × g for 10 minutes at 4°C and stored at−80°C until proteomic analysis. BALF and plasma samples from healthy volunteers enrolled in studies Pro00088966 and Pro00100375 at Duke University were also collected. BALF from these volunteers was performed under sedation and topical anesthesia, guided by chest CT scans to select the bronchopulmonary segment of interest. Similar volumes of 90 to 120 ml saline were instilled and aspirated, discarding the first 5 ml of returned fluid.

### Cohort stratification

To assess patient heterogeneity and define stratification criteria within the Munich COVID-19 cohort, t-distributed Stochastic Neighbor Embedding (t-SNE) was performed using the Rtsne package (v.0.16). The analysis was based on clinical metadata and blood chemistry parameters from 29 patients upon hospital admission. Only variables that did not require imputation at admission were included in the input matrix. To establish a longitudinal framework, clinical markers were profiled over time, including anti-SARS-CoV-2 N and S antibodies, CRP, creatinine (Jaffe method), lymphocyte counts, and leukocyte counts. Based on these profiles, three main time points were selected: day 11, day 25, and day 36. Serum aliquots from individual patients were pooled across four time windows: 0–11 days, 12–25 days, 26–36 days, and 36–54 days post-admission. The final COVID-19 cohort included 23 patients, of whom 16 were classified as severe (ICU-admitted) and 7 as mild, based on the t-SNE clustering (Table S8). As controls, serum samples were collected from patients admitted to the University Hospital of LMU Munich with suspected SARS-CoV-2 infection but who tested negative by PCR. These control patients were stratified based on systemic inflammation markers—CRP, creatinine, and leukocyte counts. Those within the third and fourth quartiles of the distribution for at least one parameter were categorized as the high-inflammatory control group, whereas those within the first and second quartiles were classified as the low-inflammatory control group. All analyses were performed using the pheatmap (v.1.0.12) and base R packages.

### Human patient material processing

Lung tissue was selected as the source for antigen profiling. A pooled lysate was generated from peritumoral lung tissue collected from 32 donors undergoing lung resection surgery for carcinoma (Table S11). All tissue samples were obtained post-transplantation and preserved under standardized conditions by the Comprehensive Pneumonology Center (CPC) BioArchive. Immediately after surgical removal, lung tissue was cut into 0.5 cm³ fragments, snap-frozen in liquid nitrogen, and stored at −80 °C until further processing. Frozen lung tissue was pulverized for protein extraction using a dry tissue pulverizer (CP02, Covaris, USA).

The resulting powder was resuspended in RIPA buffer (50 mM Tris-HCl pH 7.4, 150 mM NaCl, 1% Triton X-100, 0.5% sodium deoxycholate, 1 mM EDTA, 0.1% SDS) supplemented with protease inhibitors (Complete, Roche, Switzerland). The homogenates were incubated on ice for 30 min and then subjected to sonication using a Bioruptor device (Diagenode, Germany) for 10 cycles (30 s on, 30 s off). Following sonication, samples were centrifuged at 18,000 × g for 5 min to remove insoluble debris. Protein concentration in the cleared lysates was measured using the bicinchoninic acid assay (Pierce, Thermo Fisher Scientific, Germany).

### Differential antigen capture assay (DAC)

As previously described ^31^, immunoglobulins were then captured by incubating each sample with 20 ul of Protein L agarose beads (Pierce, Thermo Fisher Scientific, Germany). For antigen binding, a pooled lysate was prepared from peritumoral lung tissue collected from 32 age-matched individuals (Table S10). A total of 1.5 g of lung lysate was aliquoted and stored; each aliquot was thawed on ice immediately prior to use. After immunoglobulin binding, beads were incubated with the lung lysate to allow antigen–antibody complex formation. The complexes were retrieved on beads, subjected to a series of washing steps to remove nonspecific binders, and subsequently eluted for downstream mass spectrometry profiling.

### Fluid sample preparation

Plasma specimens were prepared for mass spectrometry using an automated workflow ^65^. Plasma proteins were denatured in a 96-well format, alkylated, and digested with Trypsin and LysC. Peptides were purified using an automated liquid handling platform (Agilent Bravo, USA). To construct a spectral library, 20 representative serum samples were pooled and fractionated into 24 fractions using high-pH reversed-phase liquid chromatography. This spectral library served as a reference for peptide identification in the experimental samples. BALF specimens were denatured in 4% SDS in Tris-HCl solution at 99 °C and further processed using the Protein Aggregation Capture protocol ^66^. Following digestion, peptides were purified using stage-tipping with triple-layered Octadecyl (C18)-bonded silica disks (88). Final peptide eluates were stored at −20 °C until subjected to mass spectrometry.

### Mass spectrometry

Peptides were separated using nanoflow reversed-phase chromatography on an Evosep One liquid chromatography system (Evosep). Separation was performed on an 8 cm × 150 μm column packed with 1.9 um ReproSil-Pur C18-AQ particles (Dr. Maisch). Two acquisition methods were applied depending on the experimental design. For general proteomics profiling, peptides were analyzed on a timsTOF Pro2 mass spectrometer (Bruker Daltonics) operated in data-dependent acquisition parallel accumulation–serial fragmentation (DDA-PASEF) mode over a 21 min gradient (Evosep 60 samples-per-day (SPD) method). Each acquisition cycle included three PASEF scans, with accumulation and ramp times set to 100 ms each. Singly charged precursors were excluded from acquisition, the target intensity was set to 15,000, and dynamic exclusion was applied for 0.4 minutes. Quadrupole isolation widths were set to 2 Th for m/z < 700 and 3 Th for m/z > 800. The DAC samples were analyzed using the same LC system connected to a timsTOF HT operated in data-independent acquisition parallel accumulation–serial fragmentation (DIA-PASEF) mode over a 44 min gradient (Evosep 30 samples-per-day (SPD) method). The full scan range was set from 100 to 1,700 m/z, with a ramp time of 100 ms. Ion mobility was set between 0.7 and 1.45 1/K₀. Each acquisition cycle consisted of 27 diaPASEF windows followed by one full scan, acquired across 9 frames. The DIA windows ranged from 300 to 1,110 m/z, with an isolation window of 30 Da. Collision energy was set dynamically.

### Proteomics data processing

Mass spectrometry raw files from data-dependent acquisition were analyzed using MaxQuant (v1.6.17.0) with the UniProt human reference proteome (UP000005640, version 2022_01). Searches were performed against the human and contaminant databases using the Andromeda search engine, applying carbamidomethylation of cysteine as a fixed modification and methionine oxidation and N-terminal acetylation as variable modifications. Peptide and protein identification was controlled at a 1% false discovery rate (FDR), estimated using a reverse decoy database. Trypsin and LysC were defined as cleavage enzymes, allowing up to two missed cleavages. A minimum peptide length of 7 amino acids and at least one peptide per protein group were required for quantification. Precursor mass tolerance was set to 20 ppm. DAC-based raw files acquired in data-independent acquisition mode were processed with DIA-NN (v1.8.1) in library-free mode with match-between-runs enabled. Trypsin/P and LysC were used as digestion enzymes, and methionine oxidation and N-terminal acetylation were set as variable modifications. Filtering was applied at 1% FDR for precursors and gene groups, and 0.5% for the spectral library. Protein group outputs from MaxQuant and DIA-NN were filtered to retain proteins detected in at least 3 samples per group, normalized, and imputed using a normal distribution within the DEP package (v1.24.0).

### Co-expression network analysis

Gene co-expression network analysis was applied to identify protein modules with coordinated expression patterns within each pneumonia group and characterize their functions. The analysis was conducted in R (v4.3.0) using the CoCena2 package (https://github.com/MarieOestreich/hCoCena), with log₂-transformed, normalized, and imputed protein intensity matrices from plasma and BALF samples as input (^67^. Correlation cut-offs were optimized using a weighted sum approach based on Multicriteria Decision Aiding, balancing maximization of the correlation coefficient (*R²*) and number of edges/nodes while minimizing the number of independent networks ^68^. The resulting thresholds were 0.81 for BALF and 0.74 for plasma. The network results were clustered using the Louvain algorithm ^69^ with a minimum cluster size of 10 nodes. The results were presented as a mean fold change for each module and condition in a heatmap with the module/condition setting.

### Over-representation analysis (ORA)

ORA was employed for each CoCena^2^ module to detect associated biological pathways. The analysis was performed using the clusterProfiler package (v.4.10.0) ^70^, with Reactome serving as the reference database for pathway annotation (^71^. For each module, the top 5 significantly enriched terms were selected, requiring a minimum of 5 detected proteins per term and an adjusted *p*-value < 0.05. Results were visualized as grouped dot plots, highlighting functional enrichment across modules

### Gene set variation analysis (GSVA)

GSVA was performed on imputed, log₂-transformed protein intensity matrices to evaluate longitudinal pathway behavior and molecular term dynamics ^72^. Gene sets utilized for analysis included enriched Reactome pathways identified via ORA of BALF samples, along with custom-defined gene lists. Enrichment scores were computed for individual samples using the gsva R package (v1.5.0), with a minimum gene set size of 5.

### Analysis of autoreactivity data

DAC-based proteomic data were analyzed independently for each cohort. Protein intensity values were log₂-transformed to stabilize variance. For control groups, missing values were imputed only when fewer than three non-missing values per condition were available; no imputation was performed for COVID-19 samples to avoid artificially enhancing disease-specific signals. Proteins detected in ≥3 COVID-19 patients and significantly enriched relative to at least one control group were defined as putative autoantigens [52]. COVID-19 patients were analyzed across 4 time windows, each corresponding to the next available sample following the day of intubation. In contrast, bacterial pneumonia and influenza cohorts had only 2 time points. Control groups included high-inflammatory and low-inflammatory patients from the Munich cohort, and non-pneumonia controls from the Chicago cohort.

Differential protein abundance was assessed using Welch’s *t*-test, conducted separately at each time point. Proteins with fold change >1.5 and *p* < 0.05 were considered significantly enriched. To mitigate false positives, biological relevance was evaluated based on recurrence across patients and cohorts. Significant proteins were aggregated across time points and conditions to define cohort-specific autoantigen lists.

Overlaps between cohorts were visualized using Euler diagrams generated with the Eulerr R package (v7.0.0). All analyses were performed using the Perseus software platform (v1.6.14.0) ^73^, and visualizations were generated using the ggplot2 package (v3.4.4).

### Modeling time and severity effects on autoreactivity

To investigate how disease severity and time influence autoreactivity, we analyzed longitudinal proteomic data from 23 patients in the Munich cohort, comprising 1,210 log₂-transformed protein features. Patients were grouped into *mild* and *severe* categories based on t-SNE analysis of clinical and molecular profiles. A linear mixed-effects model was applied to evaluate protein expression dynamics across four time points. The model was defined as follows:

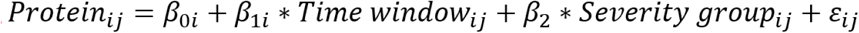

where *Protein_ij_* is the log2-transformed fold change between COVID-19 and the average low inflammatory control group of expression value for sample *i* in patient *j* for a single protein. Β_oi_ is a random intercept for the *i*th patient, β_1i_ is a random slope for the *i*th patient and β_2_ is a fixed coefficient for the severity group*. Time window_ij_* and *Severity group_ij_* represent the fixed effect for the time in the *j*th observation.

Model fitting and evaluation were performed using the lme4 (v1.1-35.1), lmerTest (v3.1-3), and psych (v2.3.9) packages. Model performance was assessed using the performance package (v0.10.8), focusing on Akaike Information Criterion, Bayesian Information Criterion, Bayes Factor, and residual normality. Significance was determined using ANOVA F-tests on fixed effects, with FDR-adjusted *p*-values < 0.05 considered significant.

### Association analysis

To investigate potential associations between autoantigen presence and clinical outcomes in severe COVID-19 patients, we performed Wilcoxon signed-rank tests. Patients positive for each autoantigen were compared to those negative for the same antigen. Results were visualized as a heatmap, with significant associations (*p* < 0.05) highlighted in red. Tests were omitted and marked in dark grey when either group had fewer than three observations (*n* < 3) or when autoantigen-positive cases were insufficient for statistical analysis. All analyses were performed in R (v4.3.2) using the ggpubr (v0.6.0), stats (v3.6.2), ggplot2, and pheatmap packages.

### Single-cell RNA-seq data analysis

Single-cell multi-omics data of peripheral blood immune cells from hospitalized COVID-19 patients and healthy controls were retrieved from ArrayExpress (E-MTAB-10026) in the.h5ad format. The dataset contained transcriptomic raw counts, surface protein data, and immune receptor sequences ^24^. LPS-stimulated and hospitalized non-COVID-19 patients were excluded from the original object using the Scanpy (v.1.9.1) package, resulting in 102 COVID-19 patients.

Subsequent preprocessing and analysis were performed in Seurat (v4.2.1). Cells with >10% mitochondrial content or <1,000 detected genes were filtered using *subset()*. Data were normalized with *NormalizeData()* using the “LogNormalize” method (scale factor = 10,000) and scaled via *ScaleData()*. Highly variable genes (HVGs) were identified using *scib.preprocessing.hvg_batch()* from scvi-tools (v0.6.8), with patient ID as the batch factor and a 15% target gene fraction. Genes with zero expression were excluded. Harmony (v0.1.1) was used to correct principal components (PCs) for patient-specific batch effects. UMAP embeddings and the neighborhood graph were computed, and clustering was performed using the Louvain algorithm with a resolution of 0.5 and 30 PCs.

Differentially expressed genes (DEGs) for initial clustering were computed using the MAST algorithm ^74^. Clusters were annotated using canonical PBMC markers ^75^. B and plasma cells were identified based on expression of marker genes (*CD79A*, *MS4A1*, *CD19*, *CD22*, *JCHAIN*, *IGKC*), and multiple IGH/IGA subclasses (*IGHG1*, *IGHG2*, *IGHG3*, *IGHG4*, *IGHA1*), and subsetted for further analysis. HVGs were recalculated for the subset, followed by data scaling, PCA, and batch correction using Harmony.

### B and plasma cell annotation

UMAP embeddings were recalculated using 20 PCs, a resolution of 0.5, and Harmony for batch correction to refine B and plasma cell subset separation. DEGs were identified per cluster using Seurat’s *FindAllMarkers()* function with the MAST algorithm ^74^. Significant DEGs were defined by a log fold change > 0.25, expression in ≥25% of cells, and Bonferroni-adjusted *p* < 0.05. The top 20 DEGs per cluster were used for manual annotation based on known B and plasma cell markers from the literature ^76^. The CITE-seq data were normalized using centered log ratio normalization via the *NormalizeData()* function^77^. Surface protein markers were identified using *FindAllMarkers()* (MAST algorithm) applied to the protein assay. Cluster annotations were refined using known markers from Woodruff et al. ^25^ and B1 cell-specific markers ^78^.

### Single-cell BCR-seq analysis

The single-cell V(D)J dataset from ^24^ were retrieved from ArrayExpress (E-MTAB-10026) and were processed following the *Single-cell Best Practices* guidelines ^79^. Filtered contig annotations were analyzed using Scirpy (v0.11.2) in Python (v3.10), retaining only cells with a single, complete B cell receptor (BCR), and excluding those with incomplete or multiple chains. Filtered contigs were merged with the single-cell B and plasma cell object using *ir.pp.merge_with_ir()*, excluding IR-negative cells, resulting in 38,006 BCR-positive cells. Clonotypes were assigned using Dandelion (v0.3.0), which accounts for somatic hypermutation by comparing all possible synonymous 5-mer variants. A distance threshold for clonotype assignment was determined using *ddl.pp.calculate_threshold()* based on the bimodal distribution of sequence similarities. Clonotyping was performed across the entire dataset. Downstream analyses, including clonotype frequency by disease severity and gene segment usage, were conducted using Scirpy and Seurat (v4.2.1).

### Study approval

The Chicago cohort study received ethical approval from the Northwestern University Institutional Review Board, and informed consent was obtained from all participants or their legal representatives. The Munich cohort study protocol and anonymized data analysis were approved by the Ethics Committee of LMU Munich (reference number 21-0047). All procedures complied with the ethical principles of the World Medical Association Declaration of Helsinki and the U.S. Department of Health and Human Services Belmont Report.

### Data availability

Raw proteomic data and MaxQuant output tables are available from the PRIDE repository^80^ under the accession numbers PXD051341 (Chicago cohort: BALF, and plasma) and PXD051982, PXD051948 (Chicago and Munich cohorts: DAC). The analysis code supporting this study is publicly available at https://github.com/schillerlab/2025_AnnaSemenova_Autoantibody.

### Statistics

Clinical metadata and clinical blood chemistry parameters were analyzed and visualized in R (v4.3.0). Group comparisons were performed using base R and the tidyverse package (v1.3.0), with visualization via ggplot2 (v3.4.4) and ggpubr (v0.6.0). Normality of continuous variables was assessed using the Shapiro-Wilk test. For non-normally distributed data, non-parametric tests were applied, including the Wilcoxon rank-sum test for two-group comparisons and the Kruskal–Wallis test for multi-group comparisons. All statistical tests were two-sided, and p-values < 0.05 were considered statistically significant.

## Author contributions

G.R.S.B., T.S.K., and H.B.S. designed the study; T.A.P. curated clinical sample collection; A.S., J.B.M.R., P.G., and M.M.H conducted experiments; A.S. and S.R.S.P. acquired and analyzed data; L.M.H., D.T., M.M., A.Ö.Y., R.G.W., A.V.M., B.D.S., G.R.S.B., and H.B.S. provided resources; A.S., and T.S.K. wrote the manuscript. All authors read and approved the manuscript.

## Funding support

H.B.S. was supported by the Helmholtz Association (Network grant CoViPa - Virological and immunological determinants of COVID-19 pathogenesis - lessons to get prepared for future pandemics), as well as the German Center for Lung Research (DZL). G.R.S.B. was supported by Simpson Querrey Lung Institute for Translational Science, the NIH (grant nos. P01AG049665, P01HL154998, U54AG079754, R01HL147575, R01HL158139, R01HL147290, R21AG075423 and U19AI135964) and the Veterans Administration (award no. I01CX001777). R.G.W. was supported by the NIH (grant nos. U19 AI135964, R01 HL149883-01, U01 TR003528, R01 AI158530, P01 HL154998) and FDA (75F40122C00134). A.V.M. was supported by the NIH (grant nos. U19AI135964, P01AG049665, P01HL154998, P01HL169188, U19AI181102, R01HL153312, R01HL158139, and R01ES034350), and research grants from AbbVie and Merck. B.D.S. is supported by NIH awards R01HL149883, R01HL153122, P01HL154998, P01AG049665, U19AI135964, and U19AI181102.

## Acknowledgements

We thank all the patients and their families for supporting the progress of science. We gratefully acknowledge the provision of human biomaterial and clinical data from the Klinikum der Universität München at Ludwig-Maximilians-Universität München. This work was supported by the German Center for Lung Research (DZL), the Helmholtz Association (CoViPa - lessons to get prepared for future pandemics) and the Max Planck Society.

## Conflict-of-interest statement

The authors have declared that no conflict of interest exists.

## Supplementary Figures

**Figure S1 - related to Figure 1.**
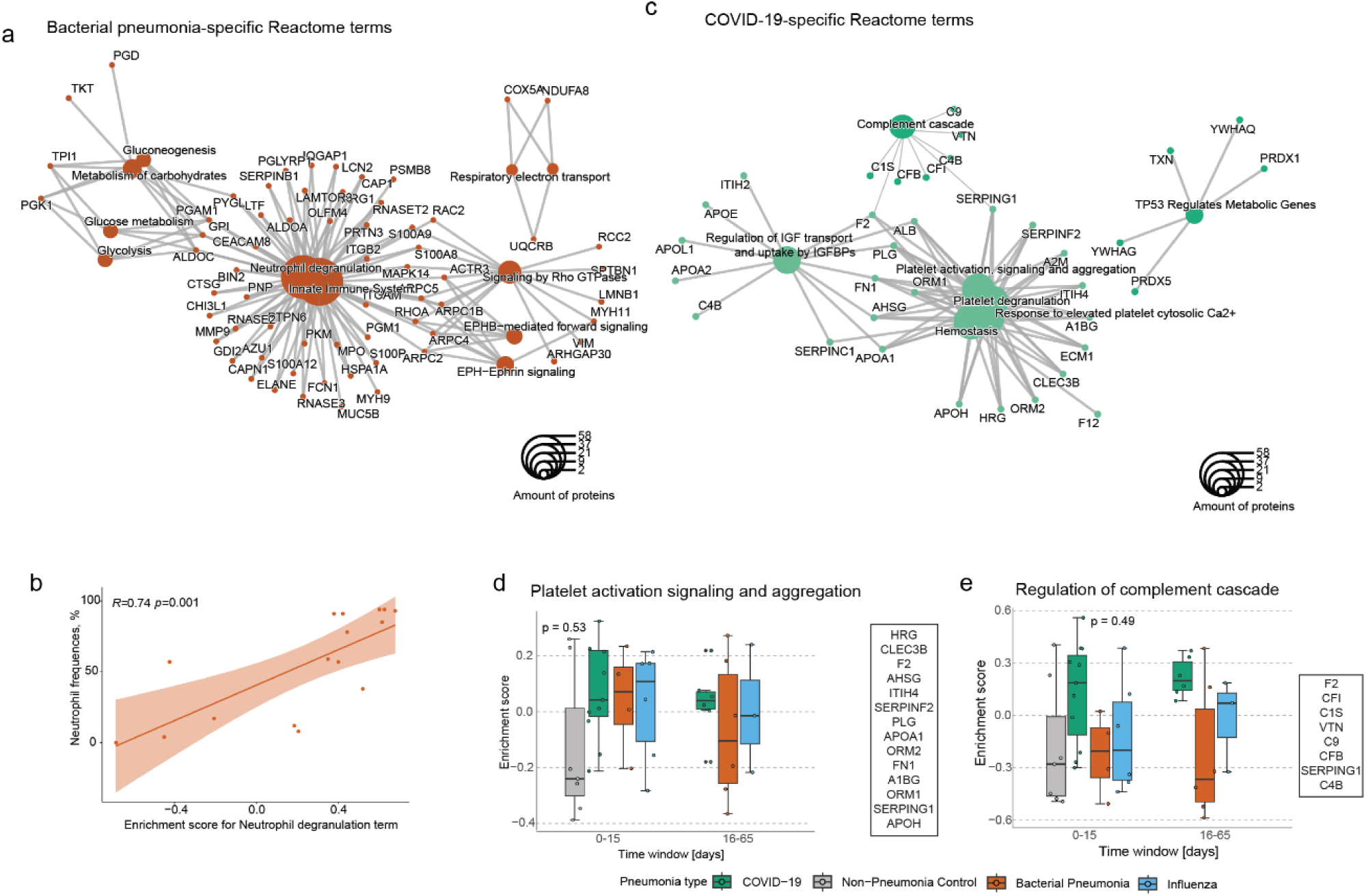
(a-b) Network plot of representative (a) bacterial-specific and (b) COVID-19-specific BALF protein modules. The circle size of the terms corresponds to the number of included proteins. (c) Pearson correlation of neutrophil degranulation proteins and normalized neutrophil frequency in the BALF of bacterial pneumonia patients. (d-e) Box plots displaying the longitudinal enrichment of (d) platelet activation signaling and aggregation and (e) regulation of complement cascade Reactome term proteins in BALF specimens across the four pneumonia types. Each dot represents the enrichment score for an individual patient. Data are represented as mean ± SD and were statistically assessed with the non-parametric Kruskal-Wallis test. Proteins involved in the term are displayed on the right side of the plot.

**Figure S2 - related to Figure 2.**
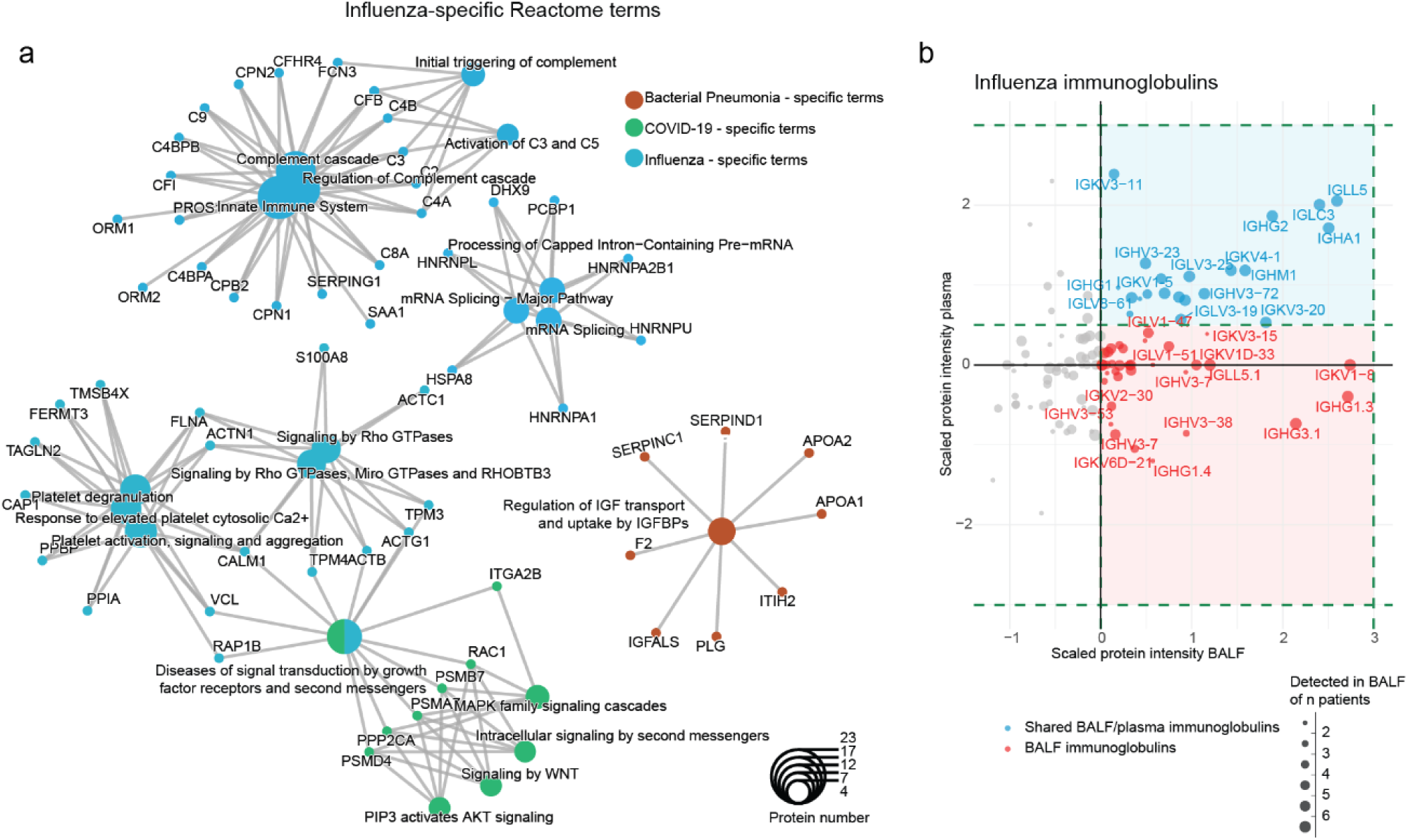
(a) Network plots of representative bacterial-specific, COVID-19-specific and influenza-specific plasma protein modules. The circle size of the terms corresponds to the number of included proteins. (b) Scatter plot displaying immunoglobulin segment abundance in the BALF (x-axis) and plasma (y-axis) of Influenza patients at the time point upon intubation. Dot size corresponds to the number of patients that express a detected protein.

**Figure S3 - related to Figure 3.**
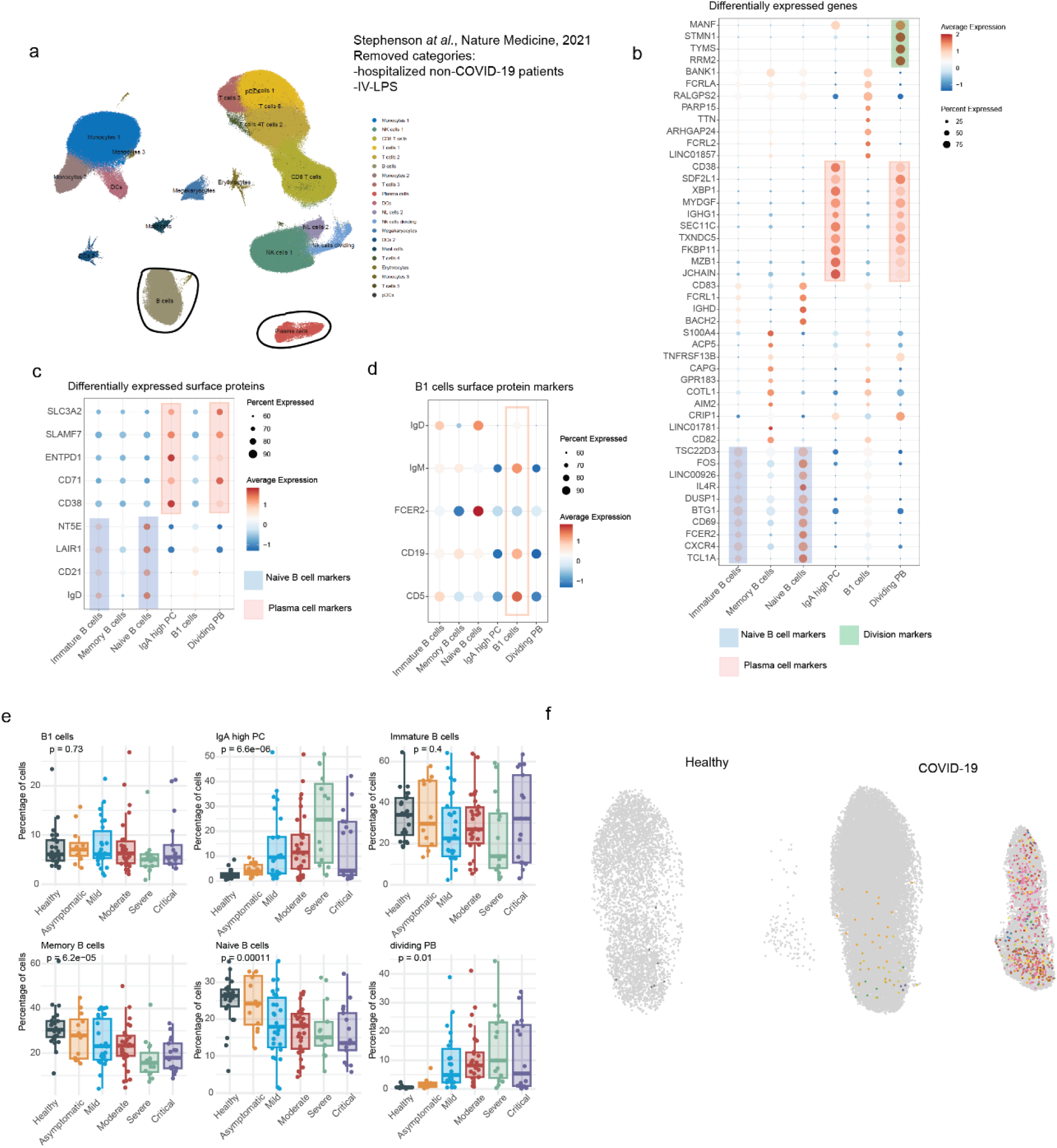
(a) UMAP plot of 632.209 PBMCs from 90 COVID-19 patients and 23 healthy controls, colored by annotation ^24^. B and plasma cell clusters are circled. (b) Dot plot of the top 10 unique gene markers for each identified B cell cluster. (c) Dot plot of differentially expressed surface protein markers for each identified B cell cluster. (d) Dot plot of B1 B cell-specific surface protein markers for each identified B cell cluster. (e) Bar plots showing the isotype distribution of B and plasma cell populations across COVID-19 disease severity groups. (f) Box plots showing the proportion of B and plasma cell populations across COVID-19 disease severity groups. Each dot represents an individual patient sample. Data are represented as mean ± SD and were statistically assessed with the non-parametric Kruskal-Wallis test. (g) UMAP visualization of 38,063 peripheral blood B/plasma cells from 90 COVID-19 patients and 23 healthy controls, colored by the expression of proteomics-identified V-segment clonotypes.

**Figure S4 - related to Figure 4.**
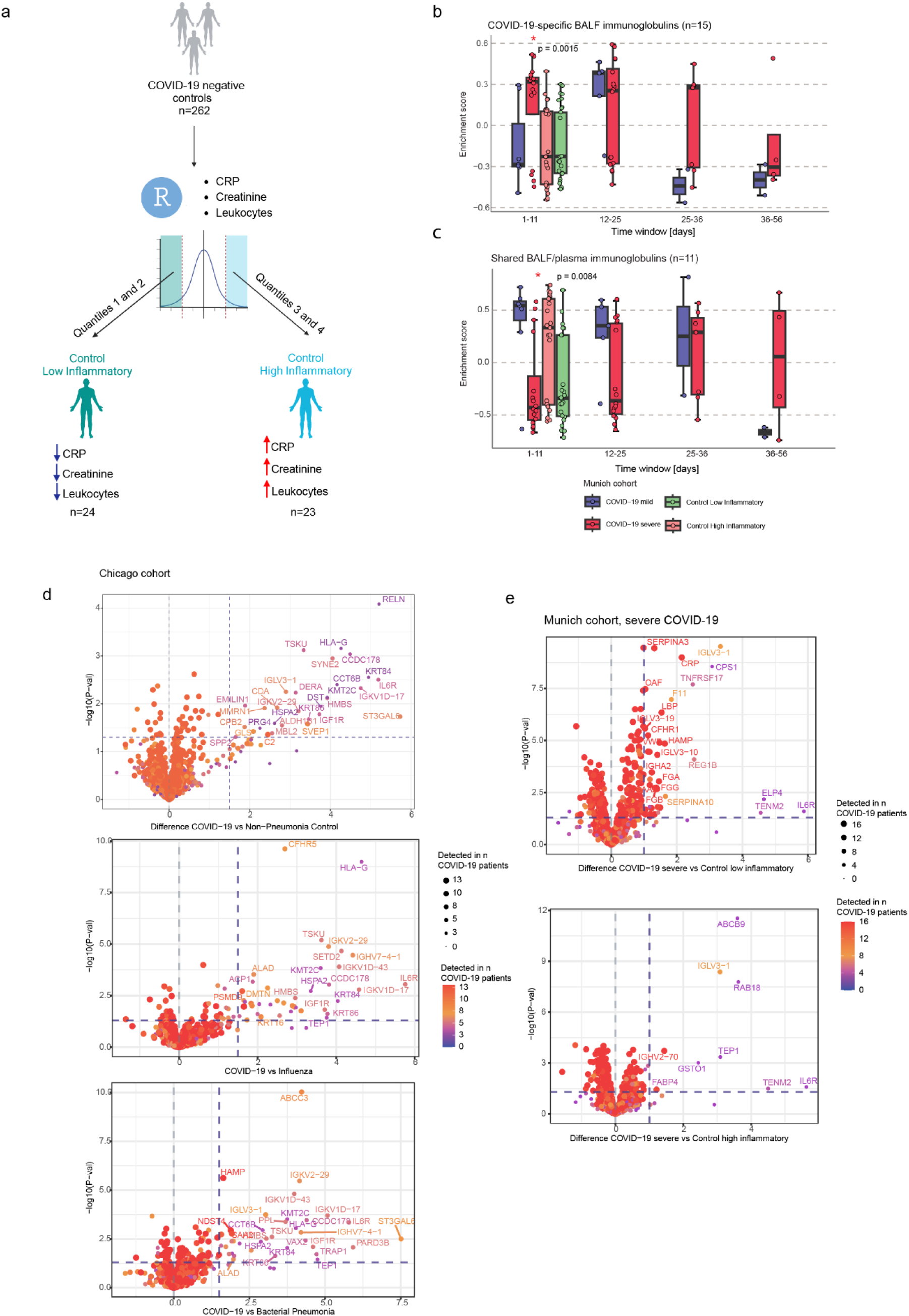
(a) Schema exhibiting the criteria of selection of control low inflammatory (n=24) and control high inflammatory (n=23) groups from SARS-CoV-2 negative patients (n=264) enrolled in the University Hospital of LMU. (b-c) Box plots displaying the longitudinal enrichment of (b) BALF and (e) shared BALF/plasma segments in BALF specimens across the four pneumonia types. Each dot represents the enrichment score for an individual patient. Data are represented as mean ± SD and were statistically assessed with the non-parametric Kruskal-Wallis test. (d-e) Volcano plots displaying immunoglobulin segment abundance between pneumonia types and controls in (d) the Chicago and (e) the Munich cohort at the time point upon intubation. Dot size corresponds to the number of patients that express a detected protein.

**Figure S5 - related to Figure 5.**
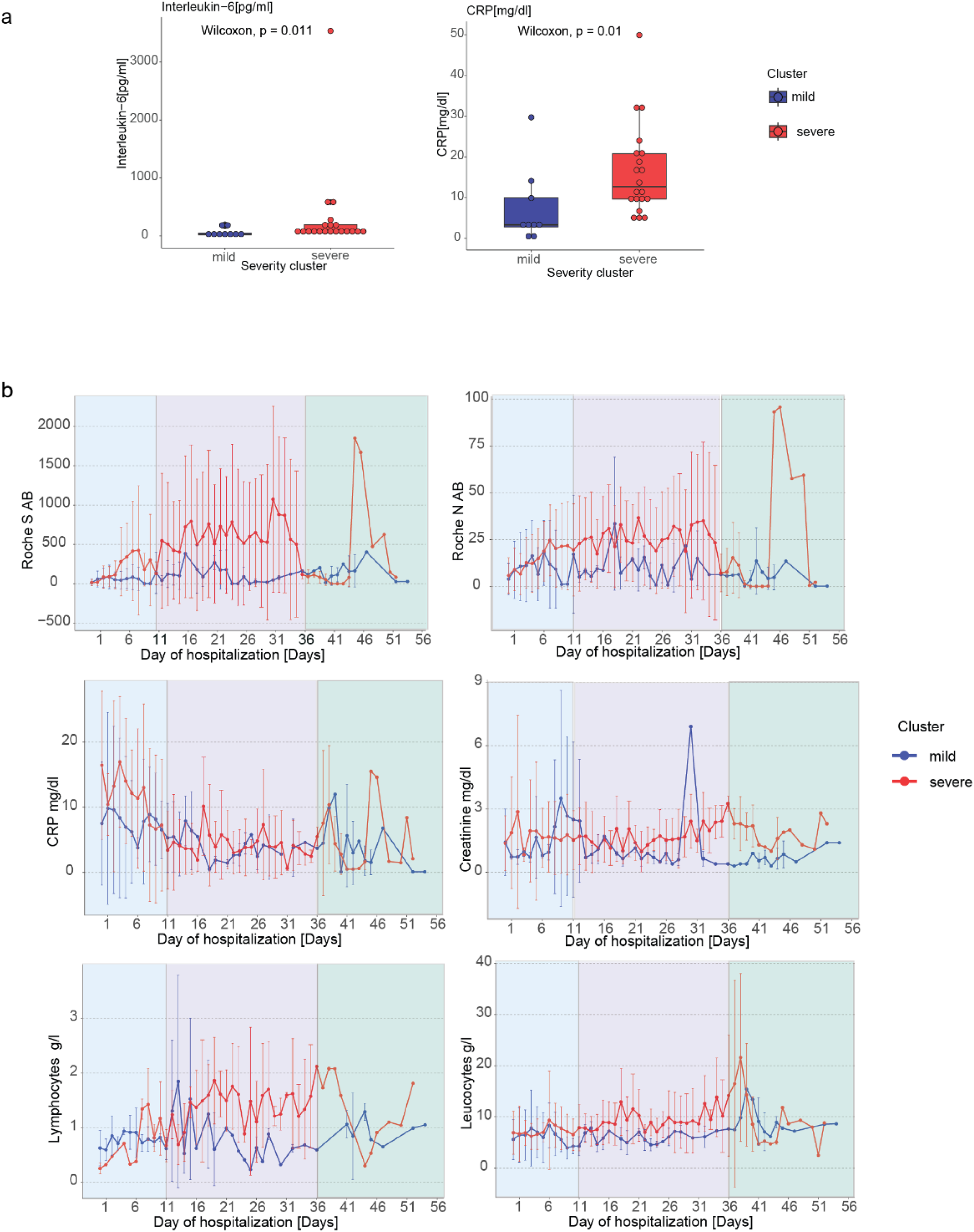
(a) Secretion levels of circulating IL-6 and C-reactive protein in the two COVID-19 patient clusters of the Munich cohort. (b) Longitudinal assessment of clinical parameters for the two COVID-19 patient clusters of the Munich cohort. Three time windows were subsequently defined: 0-11 days, 12-36 days, 37-56 days.

**Figure S6 - related to Figure 6.**
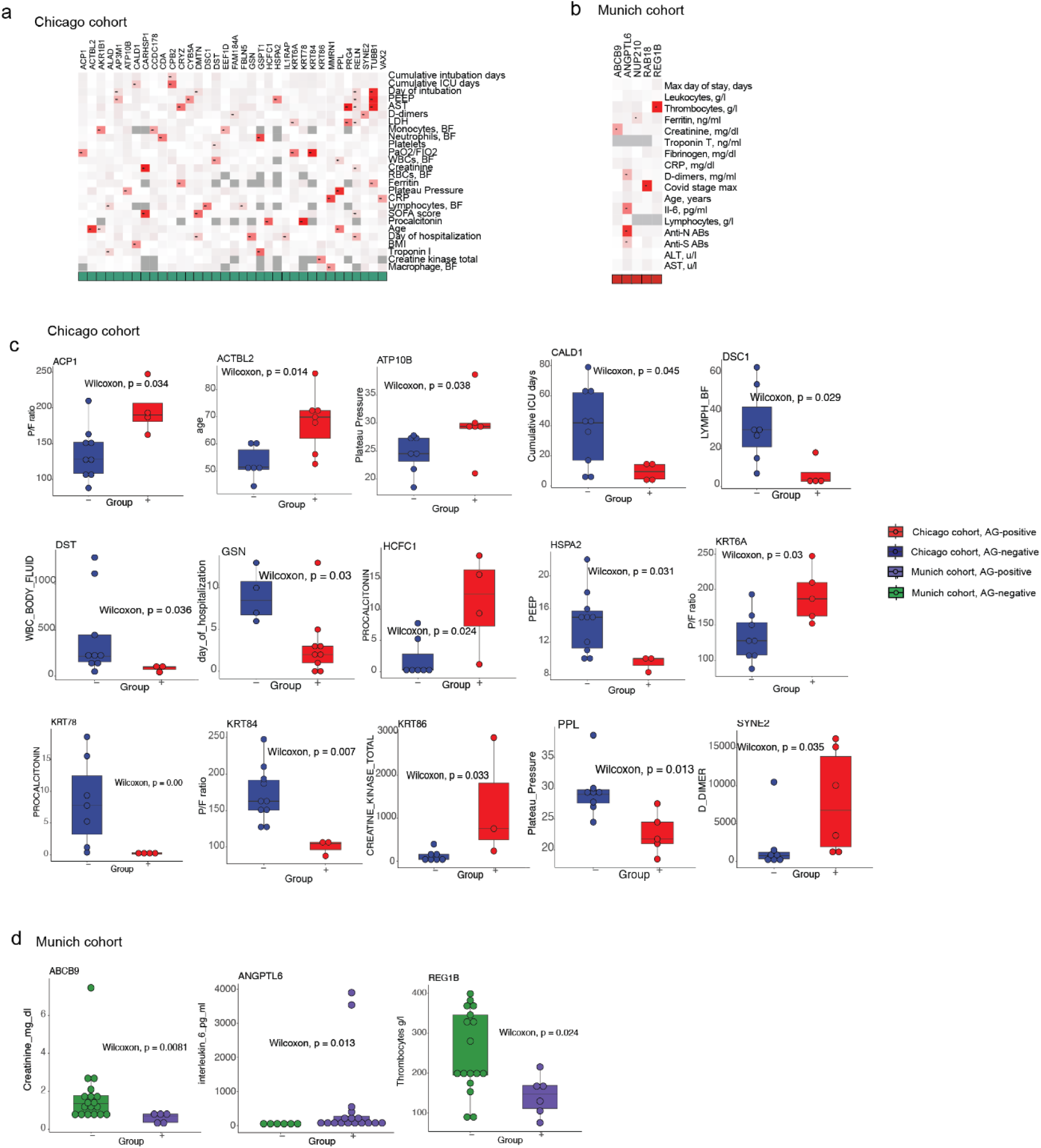
(a-b) Heatmaps presenting associations between the detection of cohort-specific putative autoantibodies in the peripheral blood of (a) COVID-19 patients (n=13) of the Chicago and (b) severe COVID-19 patients (n=16) of the Munich cohort upon intubation on the x-axis and selected clinical parameters on the y-axis. The color denotes the output of the Wilcoxon test (p-value). The dark grey color corresponds to the absence of the test results due to missing values. (c-d) Box plots showing the significant associations between putative shared autoantigens and clinical parameters in two cohorts. The x-axis represents COVID-19 patients categorized based on the presence (+) or absence (-) of detected autoantigens at the time of intubation. Statistical significance was assessed using the Wilcoxon test.

## Notes

### Competing Interest Statement

The authors have declared no competing interest.

### Author Declarations

The Chicago cohort study received ethical approval from the Northwestern University Institutional Review Board, and informed consent was obtained from all participants or their legal representatives. The Munich cohort study protocol and anonymized data analysis were approved by the Ethics Committee of LMU Munich (reference number 21‐0047). All procedures complied with the ethical principles of the World Medical Association Declaration of Helsinki and the U.S. Department of Health and Human Services Belmont Report.

